# From Wastewater to Infection Estimates: Incident COVID-19 Infections during Omicron in the U.S.

**DOI:** 10.1101/2025.09.01.25334793

**Authors:** Rachel Lobay, Ajitesh Srivastava, Daniel J. McDonald

**Affiliations:** Department of Statistics, The University of British Columbia; Department of Computer and Electrical Engineering, University of Southern California

**Keywords:** Viral Shedding, Disease burden, Reporting rate, Transmission dynamics

## Abstract

Reconstructing the course of the COVID-19 pandemic through infection estimates is important for assessing disease burden and characterizing transmission dynamics. While wastewater concentration data have been used to estimate infections in localized pre-Omicron studies, a scalable approach that incorporates reinfections and variant-specific shedding remains underdeveloped. To this end, we develop a multi-source approach to retrospectively estimate daily COVID-19 infections in U.S. states during the Omicron era. Our approach integrates wastewater and seroprevalence surveillance data and further incorporates state-specific reinfections to improve infection estimates during the Delta-Omicron transition period. These refined estimates, along with wastewater concentration data adjusted for limited coverage, are used to calculate variant-specific shedding rates, which inform daily infection estimates going forward. While case-based estimates tend to exhibit striking volatility, these infection estimates show more stable and interpretable patterns that closely align with Omicron subvariant transitions. Moreover, we directly quantify the degree of underreporting, showing the extent that reported cases significantly underestimate disease burden, with the lowest reporting rates of 9.72% in Washington, 9.73% in Minnesota, and 10.70% in New York. In the states under study, case reports capture less than a quarter of total infections, leaving the vast majority unaccounted for in official reports. Furthermore, reporting rates differ markedly across states, with disparities growing over time, reflecting the overall rise in underreporting and location-specific limitations to surveillance accuracy. Finally, we estimate time-varying effective reproduction numbers and growth rates to provide a more accurate and timely picture of transmission dynamics over the Omicron era in U.S. states.

## 1 Introduction

After the COVID-19 pandemic, reconstructing the course of infections in U.S. states is important for understanding the true timing and magnitude of outbreaks and for a more accurate assessment of disease burden. Reported cases, a convenient proxy for infections and disease burden, are lagging indicators that are subject to several delays from onset to report including initial detection, diagnostic testing, and reporting to public health (Cereda et al., 2020; Pellis et al., 2021; Zhang et al., 2020). Such delays pose major challenges for real-time estimates, which suffer from additional data quality and quantity issues, such as revision and right-censoring (Abbott et al., 2020; Arvisais-Anhalt et al., 2021; Cori et al., 2017; Linton et al., 2020; Reinhart et al., 2021). In contrast, infection estimates that are based on finalized case data (no longer subject to revision) provide a more stable and reliable reflection of disease burden over time.

Compared to finalized case data, wastewater surveillance data is a unique and underutilized alternative. It has several advantages over traditional data sources that rely on continuous mass testing efforts, such as case reports and large-scale infection prevalence studies (Chen et al., 2024a; Justman et al., 2024; Polo et al., 2020; Riley et al., 2021). In contrast to case data and prevalence surveys, wastewater data typically provides an earlier, less invasive, and more cost-effective infection indicator (Larsen and Wigginton, 2020). Moreover, studies have found reasonably strong and often sustained correlations between reported cases and viral concentrations in wastewater (Bonanno Ferraro et al., 2021; Greenwald et al., 2021; Hopkins et al., 2023; Medema et al., 2020; Rabe et al., 2023; Schill et al., 2023). As wastewater surveillance operates independently of case reporting, it may be used when testing is inadequate or reporting is delayed or no longer being reliably conducted (Larsen and Wigginton, 2020; McManus et al., 2023). In addition to allowing for more continuous monitoring, it has the potential for greater geographic coverage, especially in areas with limited testing or increased reliance on at-home testing. Furthermore, it has been shown to outperform cases in capturing patterns in COVID-19 infections during periods of high test positivity (Fernandez-Cassi et al., 2021), when compared to those from a compartmental model benchmark.

Compartmental models using wastewater concentrations have been used to to estimate COVID-19 infections for three sewersheds in South Carolina (McMahan et al., 2021) and three cities in Canada (Nourbakhsh et al., 2022). However, these models, while complex and requiring several inputs, do not account for reinfections or different variants, and so they are limited to early and short stages of the pandemic. Moreover, some of the inputs can be challenging to obtain such as the asymptomatic proportion, infectious duration, and the relative infectiousness of asymptomatic in comparison to symptomatic individuals (Nourbakhsh et al., 2022). There is the additional concern that such factors are unlikely to be time and location invariant and so should not be treated as fixed. Shedding-model-based methods (Ahmed et al., 2020, 2021) are generally simpler and more direct because they consider a multiplicative relationship between viral concentration and shedding profiles to estimate infections (Ahmed et al., 2020, 2021; Chen et al., 2024a). These methods are able to be modified using deconvolution to account for factors like delays between infection and case confirmation. However, the existing studies are on the pre-Omicron period or highly localized geographically such as those conducted on three Swiss wastewater treatment facilities (Fernandez-Cassi et al., 2021), the city of Takamatsu in Japan (Okada and Nishiura, 2024), the municipalities of Zurich and San Jose (Huisman et al., 2022), and on five counties in California (Ravuri et al., 2025). Each of these studies has a limited geographic or temporal scope and so provides limited insight into disease burden at a larger scale or during the more transmissible Omicron era. A scalable and systematic shedding-based method to estimate infections over the Omicron era remains underdeveloped.

In this study, we substantially broaden the scope by developing a novel shedding-rate and deconvolution-based methodology to estimate daily COVID-19 infections from wastewater in U.S. states, focusing on the Omicron period. We apply this to a sample of seven states, estimating daily infections from May 1, 2022 (post-BA.1) to February 1, 2023. Our analysis focuses on these states because they have both publicly available reinfection and wastewater concentration data until the end date, but the approach is broadly applicable to any state with similar data availability. To achieve this goal, we use Biobot Analytics SARS-CoV-2 wastewater concentration data (Biobot Analytics Inc., 2020), which provides extensive coverage into the Omicron period, with county and regional estimates available through 2023 (refer to Section A.1).

Figure 1 shows the data sources and steps we take to estimate infections during the Omicron period. In contrast to other wastewater-based approaches that require a number of inputs that are challenging to obtain and validate, such as the relative infectiousness, latent duration, and fraction of asymptomatic individuals (Nourbakhsh et al., 2022), our approach relies on three main data inputs: wastewater concentrations, transition period infection estimates, and variant proportions. These are leveraged to estimate shedding rates and inverse reporting ratios that directly quantify underreporting. Each ratio is then used to scale deconvolved case count data, generating the final estimate of infections. Thus, instead of estimating infections directly, our approach centers on estimating the time-varying inverse reporting ratio as an intermediate. This modular framework supports estimation during periods of variant transitions and limited case-based surveillance, provided adequate variant circulation and wastewater data are available. Moreover, since wastewater concentration data is independent of clinical testing, our framework is adaptable to periods of low or inconsistent testing and case reporting.

**Figure 1.**
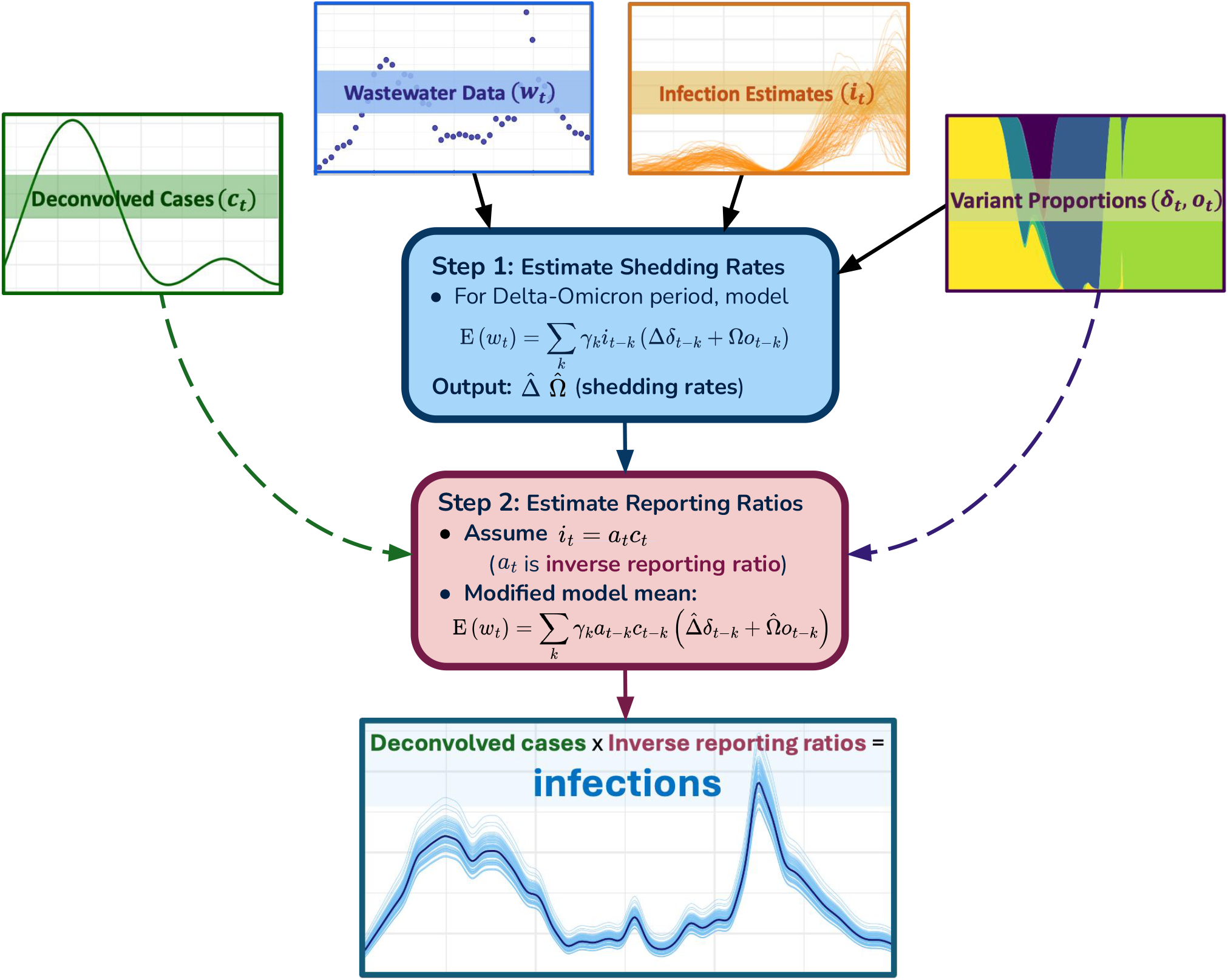
Flowchart providing an overview of the data and main steps to estimate incident infections for a state over Omicron. To provide more details on the data inputs, Infection Estimates refers to seroprevalence-based infection estimates from the Delta-Omicron transition period, adjusted for state-specific reinfections. Deconvolved Cases refers to the JHU CSSE case reports probabilistically pushed back to infection onset. Wastewater Data refers to the Biobot wastewater concentration data that has undergone a population-weighted shrinkage procedure to account for both county and regional-level data. Finally, Variant Proportions refers to the estimated proportions of Delta and Omicron variants in circulation, based on GISAID genomic sequencing data obtained from CoVariants.org (Elbe and Buckland-Merrett, 2017; Hodcroft, 2021). As for outputs, this approach produces variant-specific shedding rates and time-varying reporting ratios as intermediates in addition to the final infection estimates.

To provide an overview of each of the steps, we start with seroprevalence-based infection estimates from the Delta-Omicron overlap period, which are used in a linear model that relates infections to wastewater concentrations to derive variant-specific shedding rates. This is an important step because it allows us to estimate shedding rates for both Delta and Omicron based on the infections from the end of the Delta era—a novel contribution of this work, especially considering that previous studies do not account for differential shedding across variants (McMahan et al., 2021; Nourbakhsh et al., 2022). These estimated shedding rates, along with the variant proportions in circulation, wastewater viral concentration, and reported case data, are the primary components used to estimate the inverse reporting ratio, which represents the estimated number of infections per reported case. This ratio serves as a time-varying scaling factor for the case counts, producing the final set of infection estimates for each state. So rather than directly estimating incident infections, we first estimate this ratio as an intermediate, epidemiologically meaningful quantity that captures the change in reporting over time. This approach allows us to quantify underreporting explicitly in addition to generating estimates of infections.

The resulting infection estimates are evaluated with respect to reported cases to see how they change our understanding of the pandemic’s impact. In addition, we apply them to compute time-varying growth rates and instantaneous reproduction numbers (typically written as *R*_*t*_). These are complementary measures of transmission because effective reproduction numbers are a standard model-based indicator of transmission, while growth rates provide a more localized, model-free alternative.

This study provides a comprehensive and geographically precise approach to estimating infections that is based on wastewater concentrations, rather than case or seroprevalence survey data. This becomes increasingly important for the Omicron era, during which time traditional data sources become limited and less reliable as reinfections increase in prevalence. Our work aims to show how wastewater data can serve as a foundation to estimate infections at the state-level, providing a clearer understanding of the disease burden and improving insight into the pandemic’s impact

## 2 Methods

In this section, we describe how we estimate daily state-level infections during the Omicron period by using wastewater concentration data. To provide a conceptual understanding and motivate the approach, we first describe a simple relationship between wastewater and infections.

Suppose that, on average, the viral concentration in wastewater at time *t, w*_*t*_, reflects a signal of past incident infections *i*_*t−k*_

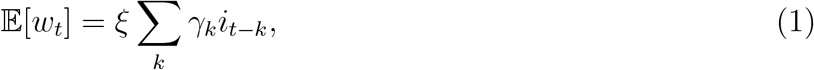

where *ξ* is a scaling factor that captures the total amount of viral material shed into wastewater by an average infection and *γ*_*k*_ is the relative fraction of total viral genetic material shed on day *k* since infection such that ∑_*k*_ *γ*_*k*_ = 1. That is, *γ*_*k*_ describes a normalized shedding profile for an individual infection over time, representing the distribution of viral shedding across days since infection, not the duration of infection. This formulation converts incidence to shedding-adjusted prevalence, computed as a weighted sum of past new infections

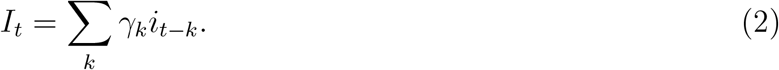

Over time, as different variants appear, *ξ* is likely to change based on the shedding characteristics of the circulating variants. To adjust for this, we extend the basic model, introducing variant-specific viral shedding rates to Eq. (1). Specifically, let Δ and Ω represent the shedding rates of Delta and Omicron, respectively, and let *δ*_*t*_ and *o*_*t*_ denote their proportions in circulation at time *t*. Then, during the period of their co-circulation, the wastewater signal is modelled as

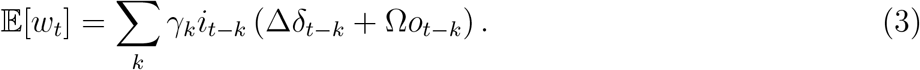

That is, the wastewater concentration reflects a smoothed sum of past incident infections, weighted by the variant-specific shedding rates and proportions in circulation. Next, we define

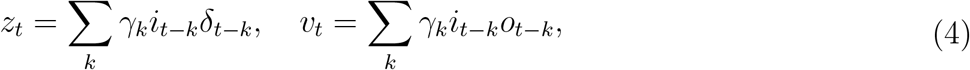

so that Eq. (3) can be expressed in the simplified form

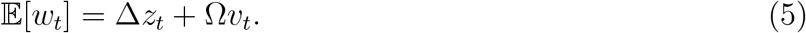

This relationship is a natural extension of the basic model, and it enables us to estimate Δ and Ω during the Delta-Omicron transition period using variant-weighted prevalence and wastewater concentration data.

To produce a forward-looking estimate of *I*_*t*_ when serology is no longer available (or is uninformative), we first examine the inverse reporting ratio, *a*_*t*_ = *i*_*t*_*/c*_*t*_, where *c*_*t*_ is the number of reported cases aligned to infection onset. Estimating *a*_*t*_ as an intermediate is informative in its own right because it directly quantifies the extent of underreporting over time.

Given estimates 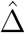 and 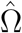 of Δ and Ω, respectively, define

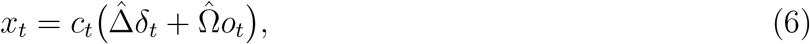

and we can model the wastewater signal as a convolution

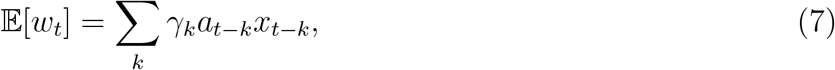

This formulation expresses *w*_*t*_ as a convolution of the inverse reporting ratio *a*_*t*_ and a known *x*_*t*_. We estimate *a*_*t*_ using deconvolution, similar to Lobay et al. (2025), but with a modified convolution matrix that includes the delay kernel *γ*_*k*_. Finally, the estimated *a*_*t*_ is used to obtain incident infections by multiplying it by the number of reported cases, which follows from the definition of the inverse reporting ratio.

In the following subsections, we provide implementation details on the two major steps of estimating the variant-specific shedding rates, 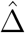, and 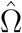, and calculating the time-varying inverse reporting ratios, *â_t_*.

### 2.1 Data processing

As shown in Figure 1, two of the main data sources used in this study are wastewater concentration data and COVID-19 case reports aligned by date of report. The raw data from both sources requires some preprocessing so that it is daily and aligned to the date of infection onset.

#### Preprocessing wastewater measurements

We use Biobot Analytics SARS-CoV-2 SARS-CoV-2 viral concentration data, normalized to the PMMoV fecal indicator to account for dilution and population size differences (Biobot Analytics Inc., 2020, 2022). This is termed effective viral concentration and is expressed in genome copies per mL of wastewater. Since this data is only provided at the county-level and the regional-level (representing many states), we construct state-level estimates by combining data from both sources. The county-level measurements within each state are reported with irregular frequencies, while the regional-level measurements are reported weekly. To construct state-level estimates for a given week, we first aggregate all available county-level observations within each state (if any), then combine these with the regional-level measurements using a convex combination based on the proportion of the state’s population accounted for in the county-level measurements. Let *p*_𝓁,*t*_ be the proportion of the state’s population covered by all county-level reports on day *t*, where 𝓁 indexes the state. Then the estimated, “shifted”, wastewater value at time *t*, denoted *w*_*t*_, is computed as

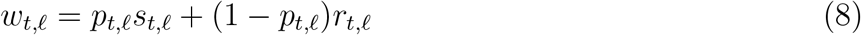

where *s*_*t*,𝓁_ is the total county-level wastewater concentration in state 𝓁 and *r*_*t*,𝓁_ is the regional wastewater concentration measurement, where the region includes state 𝓁.

The result of this shrinkage procedure is one state-level wastewater concentration estimate per state per week (see Section A.1 for more information on this). To create daily estimates for a state, we linearly interpolate these once-weekly measurements.

### Preprocessing reported case data

We use deconvolution to reconstruct the likely dates of infection onset from reported COVID-19 case data published by the JHU CSSE (Dong et al., 2020). This procedure uses estimated probabilities of the delay from infection onset to case report to “push back” aggregate cases to the estimated times of infection onset—when a person was exposed to the virus through an infected individual. Since the date a case is reported often occurs days or weeks after the person was actually infected, we use an available intermediate, namely the timing of positive specimen collection, to bridge the gap from infection to report. Therefore, this is a two-step deconvolution procedure that first propagates reported cases back to estimated specimen collection dates, and then further back to estimated infection onset. We now provide a concise summary of the two steps. For detailed methodological information, including how we estimate the delay distributions and specify the full deconvolution model, see Lobay et al. (2025).

In the first step, we estimate when each reported case likely had their positive specimen collected. While the time from infection to report is not directly observed, we use the public CDC line list that records the delay between specimen collection and reporting. By estimating probabilities that describe this delay over time, we can deconvolve reported case counts and distribute them across the earlier dates when their tests were likely collected. This step produces a smoothed time series of estimated case counts by specimen collection date, using a trend-filtering deconvolution approach. Specifically, in this nonparametric regression method, we apply an 𝓁_1_ penalty to discrete fourth-order differences in the estimated time series of case counts by specimen collection date. The 𝓁_1_ penalty results in piecewise cubic polynomials with adaptively chosen knots, enabling the model to accommodate abrupt changes in estimated cases (e.g. due to policy or behavioural changes), while still promoting smoothness. This contrasts with the global smoothness imposed by the 𝓁_2_ penalty in smoothing splines.

In the second step, we take these estimated specimen-date-aligned case counts and push them further back in time to infer when individuals were infected. Again, we base our delay distribution on empirical estimates of delays—this time from infection to specimen collection—and apply a similar deconvolution procedure that is also stratified by variant. That is, separate deconvolutions are performed for each variant category through using estimated variant proportions aligned by specimen collection date (Elbe and Buckland-Merrett, 2017; Hodcroft, 2021). The result is a set of daily infection onset estimates by variant for each state.

### 2.2 Estimating variant-specific shedding rates

To estimate infections during Omicron using wastewater data, we require a SARS-CoV-2 shedding rate for this variant. The variant-specific wastewater shedding rate refers to the total amount of viral material shed by an average infection with the variant. As shown in Eq. (3), calculating this rate specific to each of Delta and Omicron requires reliable infection estimates during the period in which they circulate. Because Delta and Omicron co-circulated during the period just before Omicron became dominant, we can estimate shedding rates for both variants if we have infection estimates from that overlap window. To this end, we use infection estimates computed by Lobay et al. (2025) that were based on seroprevalence data from two antibody prevalence surveys (Centers for Disease Control and Prevention, 2021a,c). For completeness, the model is recapitulated in Section A.2. To propagate the uncertainty in these infection estimates, we use the Gaussian process model underlying them to generate multiple plausible infection trajectories (see Section A.3).

Each set of infection estimates is used to fit the linear model given by Eq. (5). Specifically, we estimate the Delta and Omicron shedding rates by regressing the wastewater estimates on variant-weighted prevalence terms stated in Eq. (4). The data used covers the period from when the estimated prevalence of Delta or Omicron exceeds 1% in a state until February 16, 2022 (see Section A.2 for additional details on this choice). To improve estimate stability during this transition period and since the shedding rates are mainly determined by variant, impacted by factors like viral load and disease progression (Cevik et al., 2021; Hoffmann and Alsing, 2023; McMahan et al., 2021), we estimate this model jointly across states. However, it is important to note that there are some external factors (such as community demographics, the individual health profiles, and properties of treatment facilities) that may impact the shedding rates (Haver et al., 2023; Prasek et al., 2023, 2022). The complete model is is given as

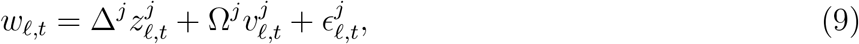

where Δ and Ω are the spatiotemporally-invariant shedding rates for Delta and Omicron that we aim to estimate, 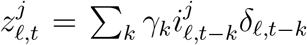 and 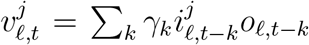 are the variant-weighted prevalence terms for Delta and Omicron, respectively and 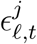 are i.i.d. Gaussian errors. This model is estimated *j* times, once for each set of infection estimates, and the resulting shedding rates are shown in Section A.4.

### 2.3 Estimating inverse reporting ratios and infections over Omicron

Given estimates of the shedding rates by variant, we return to Eq. (7) to estimate the inverse reporting ratio at each time *t* and location 𝓁, denoted as *a*_𝓁,*t*_. Importantly, this procedure uses data only after May 1, 2023, when Omicron was fully dominant, and serology measurements were no longer meaningful for tracking infections. Let *c*_*t*,𝓁_ represent the number of new deconvolved cases per 100,000 inhabitants from Section 2.1. Given variant-specific shedding rates 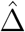 and 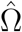, define

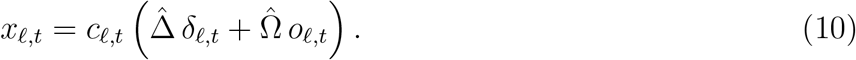

Then, we model the expected wastewater concentration at location 𝓁 at time *t* as

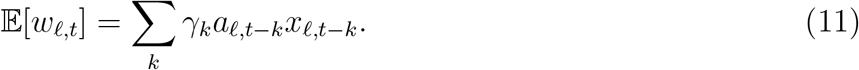

Recall that *γ* represents the normalized shedding profile distribution. Using the mean and variance for this profile from Watson et al. (2024), we apply the method of moments to fit a Gamma distribution that we subsequently discretize over a support of 0 to 17 days (Cevik et al., 2021; Russell et al., 2023), and normalize to ensure the sum of the resulting distribution is 1. Now, since we expect **a**_𝓁_ = (*a*_𝓁,1_, …, *a*_𝓁,*t*_)^T^ to be smooth in time, we estimate it using 𝓁_1_-regularized regression, which encourages local smoothness. Specifically, we solve the optimization problem

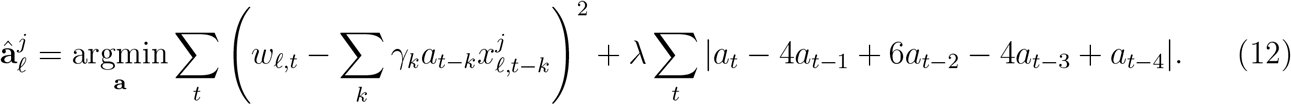

Note that the model is fit separately for each set of estimated shedding rates, indicated by the superscript *j*. The solution 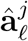 is known to be an adaptive piecewise cubic polynomial (Tibshirani, 2014, 2022) and can be accurately computed easily (Ramdas and Tibshirani, 2016). We select the tuning parameter *λ* with 3-fold cross-validation to minimize the out-of-sample error. The reason for applying the 𝓁_1_ penalty to 4^*th*^-order discrete differences in **a** is that the solution will be given as piecewise cubic polynomials (discrete splines, Tibshirani, 2022) where the number and length of each piece is chosen adaptively, allowing the model to better capture localized changes.

Each of the estimated inverse reporting ratios is then multiplied by the corresponding deconvolved case count to obtain the final infection estimate. This reporting ratio and infection estimation procedure is repeated for each of the 100 sets of shedding ratios, producing 100 sets of infection estimates for each state.

### 2.4 Time-varying instantaneous reproduction numbers and growth rates

The sets of daily infection estimates are used to compute two key epidemiological measures aimed at quantifying disease transmission dynamics over the Omicron period: the effective reproduction number, *R*_*t*_, and growth rate.

Based on our state-level infection estimates, we estimate separate *R*_*t*_ trajectories for each state. Since *R*_*t*_ represents the average number of secondary cases that result from an infected individual at a given time, producing estimates for it also requires the delay distribution describing the time between successive infections, the generation interval distribution. We estimated this for Omicron by taking the medians of the mean and standard deviation across the studies from the recent meta-analysis by Xu et al. (2023). These statistics are then matched to the shape and scale parameters of the Gamma distribution using method of moments.

To compare the *R*_*t*_ estimated from infections with that which could have been estimated from reported cases, the serial interval and incubation period distributions are estimated in the same manner as the generation interval distribution. Since the serial interval is aligned with symptom onset, a fixed shift based on the average incubation period for Omicron is applied to the resulting case-based *R*_*t*_ estimates to align them with infection onset.

*R*_*t*_ for both cases and infections is estimated by using Poisson trend filtering, meaning that the model is defined by a Poisson likelihood with a trend filtering penalty (see Liu et al. (2024)). To select the tuning parameter, 3-fold cross-validation is used to minimize the average deviance across the folds from a grid of twenty candidate values that are constructed from a logarithmically spaced grid between two bounds. The maximum bound is set as the smallest value for which the penalty term is zero once convergence and the solution have been obtained, while the minimum is set as 10^*−*4^ of that. Section A.5 shows the estimated *R*_*t*_ values for infections and cases separately, with each computed across a grid of twenty distinct tuning parameters for the state under consideration.

To assess the robustness of the infection- and case-based *R*_*t*_ estimates to the assumptions about transmission timing, we conduct a sensitivity analysis (Section A.6). In short, we examine the extreme cases: the highest and lowest average generation times for Omicron from the studies in the meta-analysis by Xu et al. (2023). If a standard deviation is unavailable, we use the same aggregated value as previously described. For the case-based *R*_*t*_, we apply the same approach using serial interval estimates, followed by a fixed incubation-period shift to better align the estimates with infection onset.

*R*_*t*_ is inherently limited by relying on modelling assumptions related to the generation time and likelihood used, amongst others. Therefore, to provide a more model-independent alternative to *R*_*t*_, we also estimate time-varying growth rates for cases and infections (Anderson et al., 2020). This measure compares the average value in two neighbouring time windows: one before and one after the reference time point. Then, the relative change between these averages is obtained and scaled by dividing by the window size to approximate the rate of change. This process is carried out over all reference time points as the window slides across the data. We start with a baseline window of size 14 to capture the relative change over consecutive two-week periods and consider alternative window sizes of 7, 21, and 28 days in a sensitivity analysis (see Section A.7).

## 3 Results

Figure 2 shows the daily estimated infections and reported cases per 100,000 population for the seven states under study. Overall, the infection trajectories for each state show two prominent waves during the summer and winter of 2022 for the transitions to the 22B and 22E subvariants. From comparing the infection and reported case curves, it is apparent that California and North Carolina tend to have closer alignment between them than the other states, especially during the major winter 2022 spike. This is numerically supported by their lower inverse reporting ratios during this time since these are the only two states that do not surpass 20 (Figure 3), suggesting more robust case surveillance and reporting systems during this period of heightened transmission. This illustrates the complementary nature of the inverse reporting ratios to the raw estimates by quantifying the extent of underreporting and visually showing its evolution over time. Furthermore, these ratios show patterns that may not be fully evident from comparing the raw infection and cases. For example, while the gap between Hawaii’s infections and reported cases appears fairly consistent across time (Figure 2), its inverse reporting ratios show a clear overall upward trend over time from below 5 to over 20 (Figure 3).

**Figure 2.**
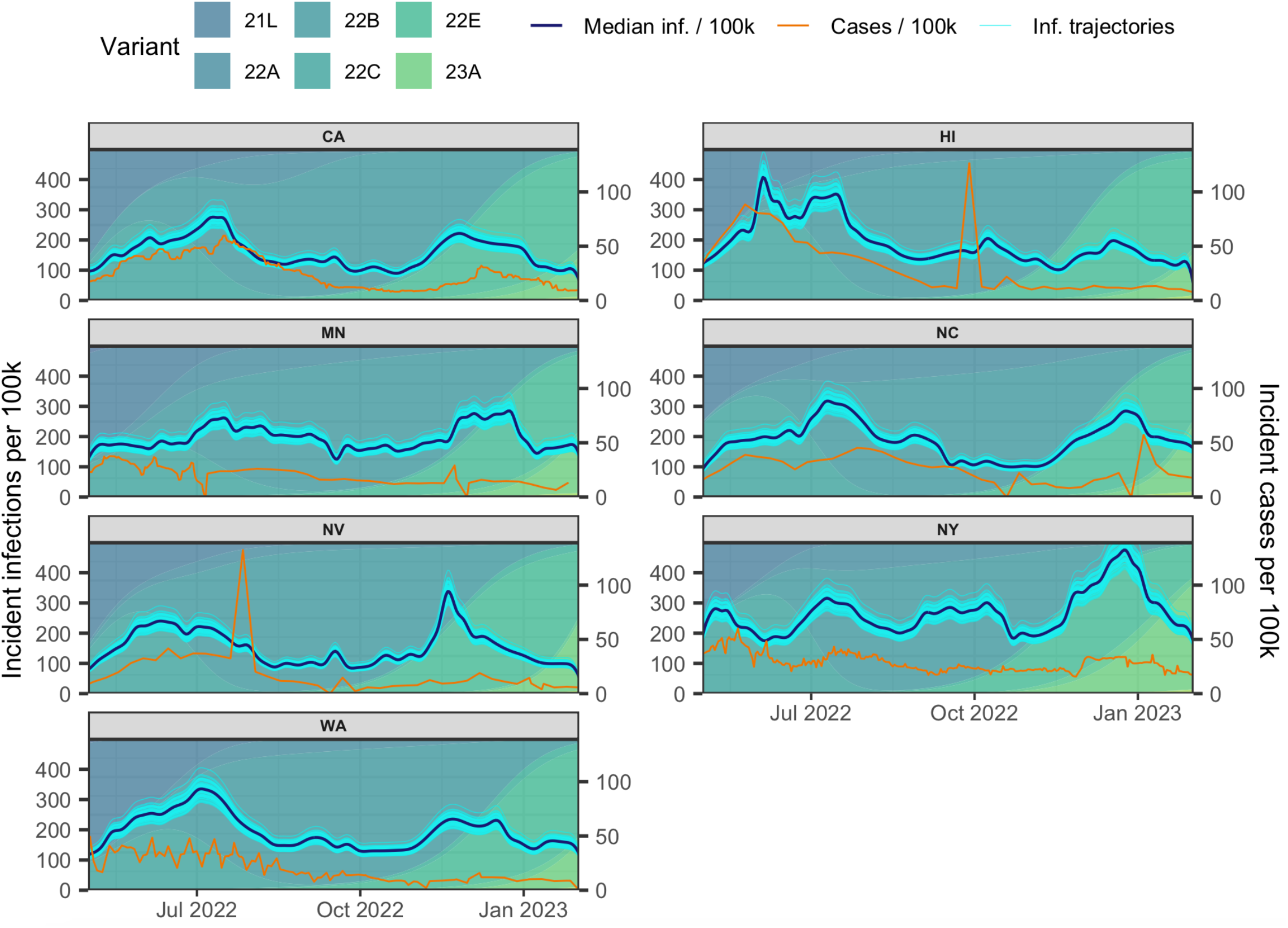
Daily estimated infections and reported cases per 100,000 population averaged over seven days for seven states. For visualization purposes, cases are transformed to match the scale of infections and the consecutive identical case values are omitted for each state, retaining only the first occurrence of each value. The background is shaded to represent the relative frequencies of Omicron subvariants in circulation, based on daily variant fractions estimated from GISAID genomic sequencing data obtained via CoVariants.org (Elbe and Buckland-Merrett, 2017; Hodcroft, 2021). This indicates the periods when a variant first appears and drives the subsequent changes in infection trends.

**Figure 3.**
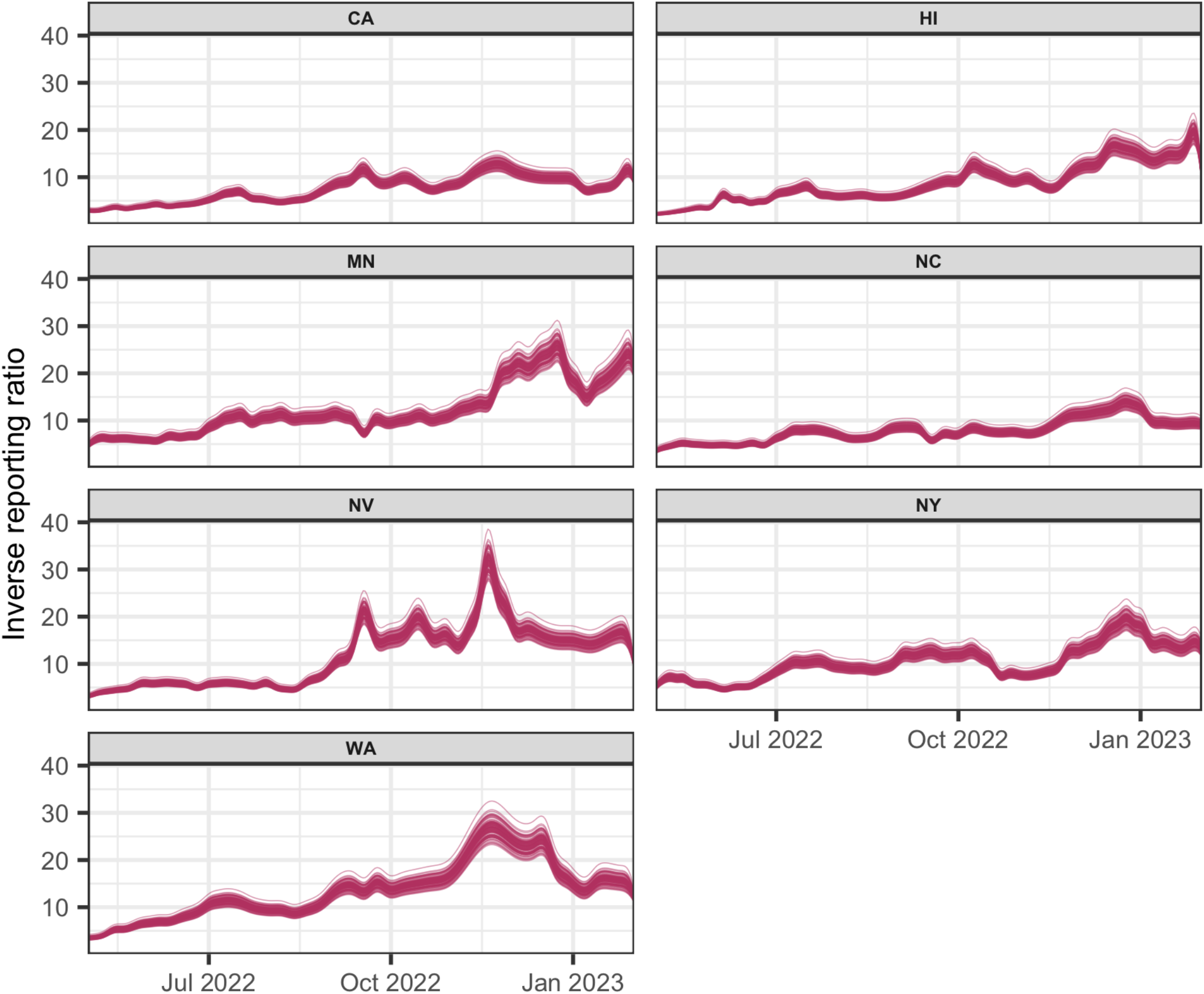
Time-varying inverse reporting ratios for seven states during the Omicron era. Note that each maroon line represents one trajectory based on a distinct set of shedding rate estimates, resulting in 100 inverse reporting ratio trajectories per state.

### 3.1 Cases versus infections and variation in underascertainment

Overall, the states show substantial and increasing underascertainment over time. Figure 2 shows that is most pronounced for New York and Nevada from mid-2022 to early 2023, though is evident in all states under study. This aligns well with the estimated infection burden as New York reaches the highest median daily infection rate of the seven states of 475 infections per 100,000 (90% quantile-based interval: 428, 524) on December 24, 2022. In contrast, its maximum case rate from around this time is only 36 per 100,000 on January 9, 2023. Following this is Hawaii with an infection rate of 407 (366, 448) per 100k on June 4, 2022 and a case rate of 89 on May 25, 2022. Next is Nevada that has a peak infection rate of 337 (304, 372) on November 19 and a corresponding case rate of 19 on December 7, 2022. Beyond these daily peaks, the cumulative estimates further reveal that reported cases substantially underestimate the actual disease burden, with the lowest reporting rates being 9.72% in Washington, 9.73% in Minnesota, 10.70% in New York, and less than 18% for all seven states under analysis. This pattern is supported by the inverse reporting ratios (Figure 3) because Washington, Minnesota, and Nevada exhibit the largest ratios overall and show a marked increase over time and peaking around 30 during the winter of 2022. The other states also attain their highest peaks in winter 2022 and their ratios exhibit upward trajectories over time, though to varying degrees.

Interestingly, the periods of prolonged underreporting have sustained spikes in wastewater-based infection estimates that are not proportionally reflected in the case data. The late-2022 surge in all seven states is the clearest example of this and underscores the utility of using wastewater data to detect such large-scale increases, especially when other surveillance sources are limited.

### 3.2 Consistent and smoother wave patterns in estimated infections

Most states present similar patterns in infections, with the two major peaks corresponding to the introduction and initial establishment of particular subvariants. This is most evident for the two major peaks that present around late June and November 2022, coinciding with the change from Omicron 22C to 22B to and 22B to 22E, respectively. In contrast, reported cases often exhibit inconsistencies and noise, which make it more challenging to identify connections to such subvariant transitions. For example, the saw-toothed pattern in the reported cases over mid-2022 from Washington (reflecting delayed and batch reporting, especially around weekends and holidays) is not evident in the estimated infections from that time, which instead show a more consistent, prolonged peak. Another example is the major mid-2022 spikes in cases in Hawaii and Nevada, which likely represent reporting backlogs because they are isolated and inconsistent with the surrounding pattern of lower infections.

### 3.3 Stability and subvariant signals in *R*_*t*_ estimates

Figure 4 shows the daily estimated infection-based and case-based *R*_*t*_ estimates for the seven states. Overall, case-based *R*_*t*_ tends to be highly volatile, which indicates the difficulty with relying on raw case data that is prone to noise and inconsistencies (due to reporting delays, periodic updates, and changes in testing practices, amongst other reasons) to derive such measures. In contrast, infection-based *R*_*t*_ lacks such instability and, hence, provides a more robust and reliable way to track transmission over time.

**Figure 4.**
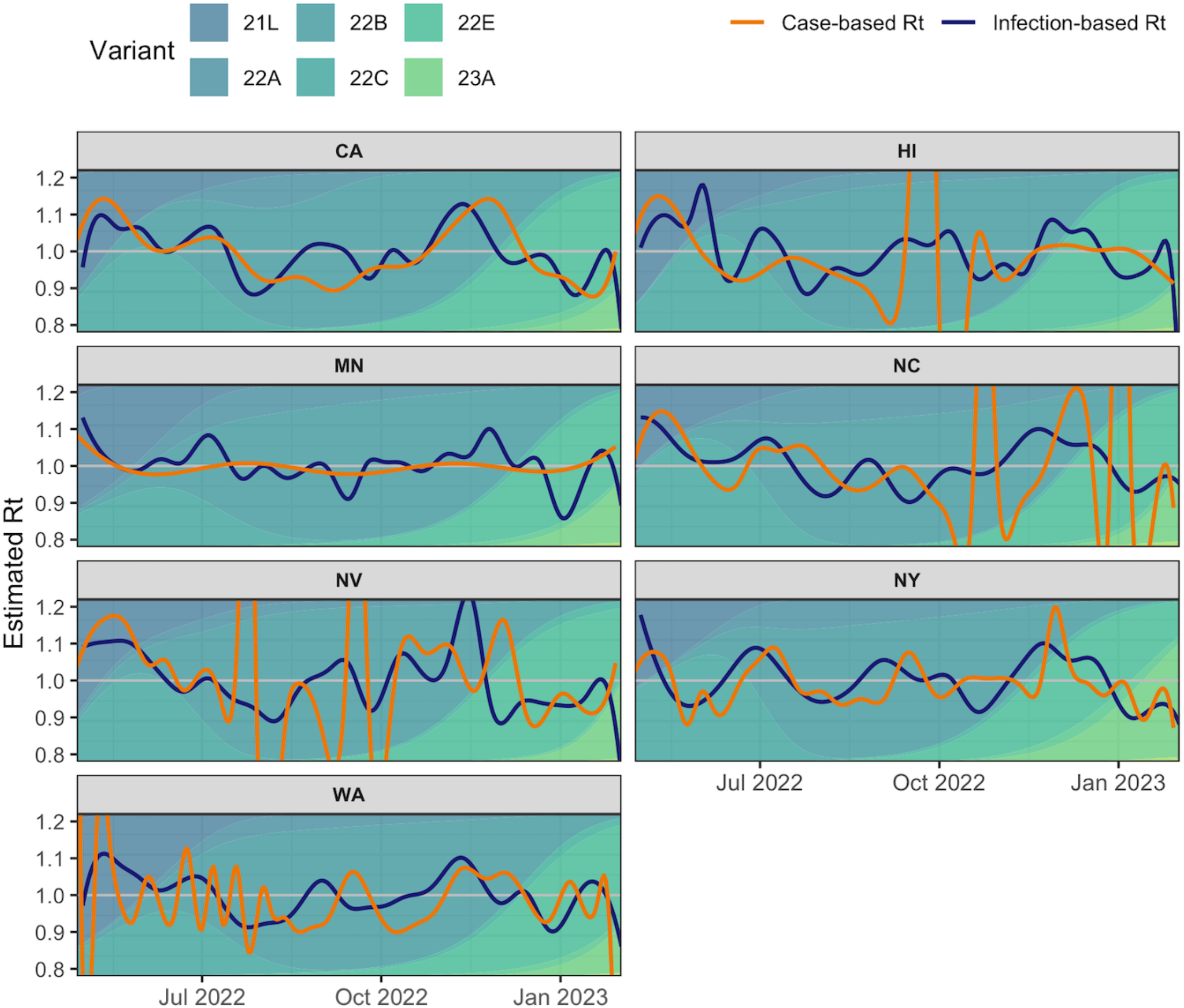
Daily estimated *R*_*t*_ using the median of infections (blue) and 7-day average of cases (orange) for the seven states of California, Hawaii, Minnesota, North Carolina, Nevada, New York, and Washington, calculated with the tuning parameter that provides the optimal cross-validation score within one standard error for each state. The background is shaded to represent the relative frequencies of Omicron subvariants in circulation, based on daily variant fractions estimated from GISAID genomic sequencing data obtained via CoVariants.org (Elbe and Buckland-Merrett, 2017; Hodcroft, 2021). This indicates the initial emergence and subsequent influence of subvariants on *R*_*t*_ trends.

Both the infection-based and case-based *R*_*t*_ fluctuate around 1, with variations in the case-based estimates strongly influenced by state-specific reporting issues and artifacts. While most states’ case-based estimates show striking volatility, Minnesota’s case-based *R*_*t*_ essentially flatlines around 1. In contrast, the infection-based estimates exhibit more pronounced oscillations, picking up on epidemic waves associated with specific subvariants (especially 22B and 23A). Given that Minnesota’s reported case counts stay consistently low with limited fluctuations (Figure 2), it is unsurprising that its case-based *R*_*t*_ fails to capture the epidemic waves as clearly as the infection-based estimates.

In general, increases in infection-based *R*_*t*_ estimates and subsequent stabilization more clearly coincide with the introduction and establishment of new variants than the case-based *R*_*t*_. For example, we observe peaks in the infection-based *R*_*t*_ during the summer and winter of 2022 that align well with the transitions to the 22B and 22E subvariants in multiple states, including New York, North Carolina, California, and Hawaii. The case-based *R*_*t*_ lack such clear patterns, exhibiting several prominent peaks likely driven by reporting artifacts, which are described in turn in Section 3.4. That said, California stands as an exception. As shown in Figure 2, its reported cases generally align well with the infection trajectories in both amplitude and timing. This results in *R*_*t*_ estimates that similarly track well and capture the same main peaks. However, its case-based signals tend to lag, especially around the peaks. During the mid-2022 wave, infections peaked on July 10, while case rates peaked six days later (and differed substantially in scale at 276 versus 54 per 100,000). A similar, though shorter, delay is reflected in the *R*_*t*_ estimates: the infection-based *R*_*t*_ peaked at 1.07 on July 4, followed by case-based *R*_*t*_ peaking at 1.04 on July 6. A longer delay is evident in the late 2022 estimates, where infection-based *R*_*t*_ peaks on November 12, while the case-based estimate lagged by 13 days. These examples show that even when infection and case trajectories align well, as in California, case-based *R*_*t*_ tends to be delayed during the period of peak infection, providing late warnings of transmission.

### 3.4 Backlogs and reporting inconsistencies in case-based *R*_*t*_

States with weekly or periodic reporting typically have more stable case-based *R*_*t*_ than states with daily reporting, but they are also more susceptible to issues like reporting backlogs. As shown in Figure 2, New York’s more consistent daily reporting cadence makes case-based *R*_*t*_ more volatile due to small day-to-day changes (which arise from factors such as weekends, holidays, reporting errors, delays in the reporting pipeline, and fluctuations in test positivity). In mid-2022, the saw-toothed pattern in Washington’s case reports leads to large-scale fluctuations in its Rt, while in late 2022, North Carolina experiences extreme spikes that coincide with sharp drops in reported cases to zero.

Similarly, the most extreme spikes in Hawaii and Nevada’s case-based *R*_*t*_ estimates do not temporally align with subvariant establishment or change-points, but instead coincide with the major backlogs observed in infections in mid-2022 (Figure 2). In contrast, the infection-based *R*_*t*_ for these states remains more stable, suggesting that actual transmission did not surge at that time, only case reporting did.

### 3.5 Volatility and reliability in growth rates

For every state under study, the case growth rates are more volatile than the infection growth rates (Figure 5). Whereas the infection-based growth rates remain relatively constrained within ± 1%, the case-based growth rates exhibit large swings indicating rapid short-term increase or decrease, respectively. For example, Nevada and Hawaii show periods of extreme increase and decrease in growth rates over fall 2022 that coincide with their reporting backlogs (also reflected in the case-based *R*_*t*_ estimates). Minnesota and North Carolina show similar extreme patterns as the drops to low or zero case rates during winter 2022 lead to volatile growth rates. In addition to these extreme patterns, there are also frequent small-scale fluctuations, most notably in Washington and New York, due to pronounced volatility in case rates (see Figure 2). Collectively, these examples support that the case-based growth rates are more susceptible to reporting artifacts, making them less reliable, especially when compared to the more stable infection-based growth rates during the same period.

**Figure 5.**
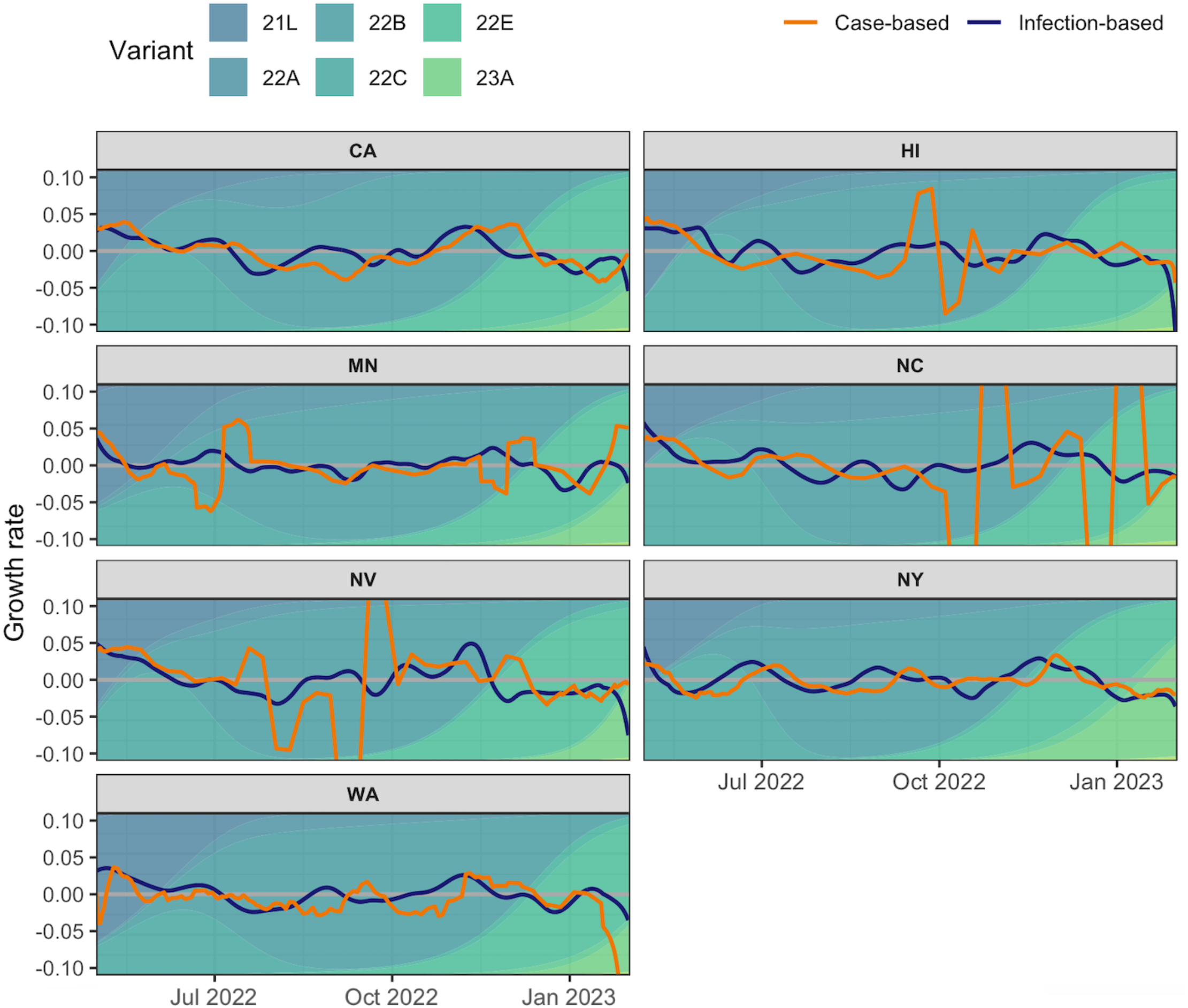
Daily relative change growth rate estimates for infections (blue) and the 7-day average of cases (orange) for the seven states of California, Hawaii, Minnesota, North Carolina, Nevada, New York, and Washington. Note that the growth rates are calculated using the data that falls within 14-days above and below the time under consideration.

To assess the robustness of the growth rate estimates to the size of the smoothing window, we conduct a sensitivity analysis using bandwidths of 7, 21, and 28 days, in addition to the default 14-day window (Section A.7). The results show qualitatively similar patterns to those described above, albeit with increased smoothness as the window size increases—especially for the case-based estimates, which are more vulnerable to noise and reporting irregularities.

## 4 Discussion

We estimate daily incident infections for a sample of seven U.S. states from May 1, 2022 to February 1, 2023. Most importantly, these wastewater-based infection estimates reveal significant underreporting during the circulation of Omicron 21L to 23A. Over this time, reported cases captured less than 18% of total infections, and Washington reported as few as 9.72%. Moreover, these infection estimates mitigate inconsistencies observed in case data, such as backlogs and irregular reporting, and better align with subvariant transitions (most notably from 22C to 22B and 22B to 22E). In addition, our methodology produces epidemiologically meaningful intermediates, namely variant-specific shedding rates and inverse reporting ratios that dynamically quantify underreporting alongside our primary output of infection estimates. We also use these reconstructed infection estimates to produce time-varying effective reproduction numbers (*R*_*t*_) and growth rates, which provide important insights into disease transmission, particularly in periods of unstable case reporting and underreporting. Overall, the infection-based *R*_*t*_ estimates are more reliable than the case-based estimates, as they better capture the emergence of new variants, while the latter are often distorted by noise and erratic fluctuations.

While wastewater concentration data is the primary source used to drive the estimation process forward, this study also incorporates case and seroprevalence data via the Delta-Omicron infection estimates. In Lobay et al. (2025), incident infections for U.S. states were estimated using case and seroprevalence data (for the reported and unreported infections, respectively). However, relying on seroprevalence data to estimate infections became increasingly unjustifiable for two reasons. First, the widespread CDC seroprevalence surveys both concluded by early 2022 (Centers for Disease Control and Prevention, 2021a,c). While the CDC conducted a 2022–2023 Blood Donor Seroprevalence Survey, only quarterly estimates are reported, so it lacks the temporal resolution needed for daily estimation (Centers for Disease Control and Prevention, 2021b). Second, after December 2021, estimating time-varying case-ascertainment with seroprevalence data alone becomes challenging because seropositivity nears 100%.

Aside from seroprevalence, incorporating location-specific and time-varying reinfection data is essential for accurately estimating COVID-19 infections. Excluding reinfections leads to an underestimation of the inverse reporting ratio and consequently infections, while misapplying them from one location or time period to another can lead to misleading infection estimates (Centers for Disease Control and Prevention, 2021d; Chen et al., 2024b; Keeling, 2023). However, even within a state, the completeness of reinfection data varies due in part to the changing definition as well as testing behaviour and reporting practices (Ruff et al., 2022; Washington State Department of Health, 2022). Moreover, this became a cross-state concern with the emergence of Omicron BA.1 as the likelihood of reinfection rose sharply and continued to rise with later subvariants (Ma, 2023).

Reinfection and other standard sources of surveillance data are subject to similar testing and reporting issues. For example, case, reinfection, hospitalization and death data are subject to underreporting, variable testing, and substantial reporting delays (Fox et al., 2022). Thus, incorporating data sources that are not subject to these same type of delays is important for deriving more comprehensive and accurate estimates of infections (Kaplan et al., 2022). In comparison to seroprevalence and the conventional sources of case, hospitalization and death data, wastewater data tends to be less invasive and costly (Larsen and Wigginton, 2020). While viral shedding tends to occur within a constrained time frame, there have been cases of viral shedding that persist for weeks or even months post-recovery and recent work has begun to investigate its impact on daily incidence (Cevik et al., 2021; Natarajan et al., 2022; Phan et al., 2025). In our modelling procedure, viral shedding is represented by the shedding profile and, hence, we were able to investigate how different empirically grounded profiles affect the final infection estimates.

In a sensitivity analysis, we considered our original choice and two alternative shedding profiles (Huisman et al., 2022; Okada and Nishiura, 2024) for which the means and standard deviations were provided and proceeded as before. This investigation was limited by the availability of shedding profiles applicable to both the temporal and spatial contexts under study. Although neither profile fully satisfies these criteria, we opted for these because of their relative strengths: the former covers part of the Omicron period, while the latter, though predating Omicron, is based on U.S. data and has been used in related studies (see, for example, Ravuri et al. (2025)). Under each of these alternative shedding profiles, the same procedure was performed as was done originally to estimate the statewise infection trajectories. The results showed similar magnitude, shape, and spread of the infections and consistency with the main results (see Section A.8). This consistency reinforces the reliability of our infection estimates despite uncertainty in the exact timing and magnitude of viral shedding in wastewater (Huisman et al., 2022). Moreover, present studies tend to assume uniformity and homogeneity in viral shedding behaviour, when it can depend on the variant type, disease severity, and other factors (Cevik et al., 2021; Chen et al., 2024a; Prasek et al., 2023). As more information on the shedding profile and its sources of variability emerges, future extensions of our approach could incorporate variant-specific or other customized shedding profiles instead of a fixed *γ* in our wastewater-centric model.

It is also important to recognize that the Biobot wastewater concentration data is not continuously available for the contributing communities over the course of the COVID-19 pandemic. While locations that are part of the Biobot Network are requested to sample weekly (Biobot Analytics Inc., 2021), infrequent submissions and retroactive updates create significant data gaps (Biobot Analytics Inc., 2023). Much of the challenge in using this wastewater data stems from these gaps. A clear example of this is seen in Figure A.2, where there is an extensive gap in wastewater estimates over the fall and winter of 2021 in North Carolina. The shrinkage procedure described in Section 2.1 helps to use regional-level data effectively to fill-in these gaps, providing a more principled approach than smoothing methods that rely on heuristically chosen parameters (e.g., moving averages and LOESS).

In addition to such time-based limitations, there are also geographic limitations that affect the reliability of wastewater concentration data. For example, county-level data relies on sewershed-restricted samples that are not entirely representative of the county, and hence, introduce bias into the wastewater concentration measurements (Ravuri et al., 2025). Moreover, measurements are typically only available for a limited number of counties in a state and, thus represent a fraction of the state. For example, North Carolina and New York had wastewater concentration data covering only 1.7% (90% quantile-based interval: 0.0%, 4.7%) and 2.3% (90% quantile-based interval: 0.0%, 6.9%) of their respective state populations on average. Washington followed with a slightly higher average coverage of 7.1% (90% quantile-based interval: 0.0%, 22.0%). All of the others had a mean coverage of at least 17.8% and Nevada had the highest mean coverage at 48.4% (90% quantile-based interval: 0.0%, 88.7%). This tendency towards low coverage limits generalizing to the state-level without additional data or supplementation. While regional data can help to capture broader trends, it lacks the resolution to reflect state-specific trends by itself. When strategically combined, county and regional data can contribute to a more accurate approximation of state-level trends than either does alone (refer to Section A.1).

Despite these limitations, wastewater concentration data is being increasingly used to complement other surveillance streams and improve COVID-19 infection estimation (Chen et al., 2024b). Since case and hospitalization data are affected by disease-related and systemic factors such as disease progression, reinfections, and testing availability, other avenues like wastewater concentrations should be considered to track infections going forward as these issues become increasingly severe (Huisman et al., 2022; Schill et al., 2023; Soller et al., 2022). Wastewater data does not depend on the availability and accessibility of testing or other limited healthcare resources such as hospital capacity. Improving its coverage and consistency, as well as further clarifying its relationship with case and hospitalization reporting, would strengthen its reliability for both retrospective and real-time estimation.

More broadly, further research is necessary to determine the extent that disease incidence can be detected and monitored by using wastewater data. Several considerations should ideally be factored in such as the disease transmission dynamics, geographic locations, population characteristics, as well as the sampling and collection procedures (Larsen and Wigginton, 2020). In addition, more research should be conducted into how to best leverage wastewater data in conjunction with standard sources of data to estimate key measures of disease frequency and to reconstruct the course of COVID-19 over time.

Overall, our work on estimating COVID-19 infections over Omicron in U.S. states is important to both the public and to public health as it is part of the broader effort to reflect on the pandemic and to advance scientific and public understanding on it. The shedding rates and reporting ratios are informative intermediates, while the resulting infection estimates can be analyzed independently or applied to derive reproduction numbers and growth rates to assess transmission over time. Moreover, our flexible methodology provides a foundation for ongoing and future investigations to assess the impact of COVID-19 according to major epidemiological and public-health metrics.

## Data Availability

All data produced are available online at

https://github.com/cmu-delphi/incident-infections-omicron/

## Data availability

The code to reproduce all figures and results is available at https://github.com/cmu-delphi/incident-infections-omicron/.

## Acknowledgements

We are grateful to members of the Delphi research group, particularly Ryan J. Tibshirani, for insightful early conversations and preceding work.

We acknowledge all data contributors, i.e., the Authors and their originating laboratories responsible for obtaining the specimens, and their submitting laboratories for generating the genetic sequence and metadata and sharing via the GISAID Initiative (Elbe and Buckland-Merrett, 2017), on which this research is based.

Any opinions, findings, and conclusions or recommendations expressed in this material are those of the authors and do not necessarily reflect the views of the National Science Foundation and the Centers for Disease Control and Prevention.

AS was supported by the Centers for Disease Control and Prevention and the National Science Foundation under Award No. 2223933 and 2333494. DJM was supported by Centers for Disease Control and Prevention (CDC) Grant No. 75D30123C15907. DJM and RL received support from the National Sciences and Engineering Research Council of Canada and the University of British Columbia.

## References

Abbott, S., Hellewell, J., Thompson, R.N., Sherratt, K., Gibbs, H.P., Bosse, N.I., Munday, J.D., Meakin, S., Doughty, E.L., Chun, J.Y., et al., 2020. Estimating the time-varying reproduction number of SARS-CoV-2 using national and subnational case counts. Wellcome Open Research 5, 112.

Ahmed, W., Angel, N., Edson, J., Bibby, K., Bivins, A., O’Brien, J.W., Choi, P.M., Kitajima, M., Simpson, S.L., Li, J., et al., 2020. First confirmed detection of SARS-CoV-2 in untreated wastewater in Australia: a proof of concept for the wastewater surveillance of COVID-19 in the community. Science of the Total Environment 728, 138764.

Ahmed, W., Tscharke, B., Bertsch, P.M., Bibby, K., Bivins, A., Choi, P., Clarke, L., Dwyer, J., Edson, J., Nguyen, T.M.H., et al., 2021. SARS-CoV-2 RNA monitoring in wastewater as a potential early warning system for COVID-19 transmission in the community: A temporal case study. Science of The Total Environment 761, 144216.

Anderson, R., Donnelly, C., Hollingsworth, D., Keeling, M., Vegvari, C., Baggaley, R., Maddren, R., 2020. Reproduction number (R) and growth rate (r) of the COVID-19 epidemic in the UK: methods of estimation, data sources, causes of heterogeneity, and use as a guide in policy formulation. The Royal Society.

Arvisais-Anhalt, S., Lehmann, C.U., Park, J.Y., Araj, E., Holcomb, M., Jamieson, A.R., McDonald, S., Medford, R.J., Perl, T.M., Toomay, S.M., et al., 2021. What the coronavirus disease 2019 (COVID-19) pandemic has reinforced: the need for accurate data. Clinical Infectious Diseases 72, 920–923.

Biobot Analytics Inc., 2020. Biobot web dashboard – COVID-19 wastewater monitoring in the us. https://biobot.io/data/covid-19.

Biobot Analytics Inc., 2021. The Biobot network opt-in. https://www.app.biobot.io/sign-up.

Biobot Analytics Inc., 2022. Effective concentration: a future-proof approach to characterizing pathogen concentrations in wastewater. https://web.archive.org/web/20230406191212/ https://biobot.io/wp-content/uploads/2022/02/2022-01-White-paper-Effective-concentration.pdf.

Biobot Analytics Inc., 2023. Biobot FAQs - dashboard. https://support.biobot.io/hc/en-us/articles/15952615508119-FAQs-Dashboard.

Bonanno Ferraro, G., Veneri, C., Mancini, P., Iaconelli, M., Suffredini, E., Bonadonna, L., Lucentini, L., Bowo-Ngandji, A., Kengne-Nde, C., Mbaga, D., et al., 2021. A state-of-the-art scoping review on SARS-CoV-2 in sewage focusing on the potential of wastewater surveillance for the monitoring of the covid-19 pandemic. Food and Environmental Virology, 1–40.

Centers for Disease Control and Prevention, 2021a. 2020-2021 nationwide blood donor seroprevalence survey infection-induced seroprevalence estimates. https://data.cdc.gov/Laboratory-Surveillance/2020-2021-Nationwide-Blood-Donor-Seroprevalence-Su/mtc3-kq6r.

Centers for Disease Control and Prevention, 2021b. 2022-2023 nationwide blood donor seroprevalence survey infection-induced seroprevalence estimates. https://data.cdc.gov/Laboratory-Surveillance/2022-2023-Nationwide-Blood-Donor-Seroprevalence-Su/ar8q-3jhn/.

Centers for Disease Control and Prevention, 2021c. Nationwide commercial labo-ratory seroprevalence survey. https://data.cdc.gov/Laboratory-Surveillance/Nationwide-Commercial-Laboratory-Seroprevalence-Su/d2tw-32xv.

Centers for Disease Control and Prevention, 2021d. Nationwide COVID-19 infection-induced antibody seroprevalence (commercial laboratories). https://covid.cdc.gov/covid-data-tracker/#national-lab.

Cereda, D., Tirani, M., Rovida, F., Demicheli, V., Ajelli, M., Poletti, P., Trentini, F., Guzzetta, G., Marziano, V., Barone, A., et al., 2020. The early phase of the COVID-19 outbreak in Lombardy, Italy. arXiv doi:10.48550/arXiv.2003.09320.

Cevik, M., Tate, M., Lloyd, O., Maraolo, A.E., Schafers, J., Ho, A., 2021. SARS-CoV-2, SARS-CoV, and MERS-CoV viral load dynamics, duration of viral shedding, and infectiousness: a systematic review and meta-analysis. The Lancet Microbe 2, e13–e22.

Chen, C., Wang, Y., Kaur, G., Adiga, A., Espinoza, B., Venkatramanan, S., Warren, A., Lewis, B., Crow, J., Singh, R., et al., 2024a. Wastewater-based epidemiology for COVID-19 surveillance and beyond: A survey. Epidemics 49, 100793.

Chen, Y., Zhu, W., Han, X., Chen, M., Li, X., Huang, H., Zhang, M., Wei, R., Zhang, H., Yang, C., et al., 2024b. How does the SARS-CoV-2 reinfection rate change over time? The global evidence from systematic review and meta-analysis. BMC Infectious Diseases 24, 339.

Cori, A., Donnelly, C.A., Dorigatti, I., Ferguson, N.M., Fraser, C., Garske, T., Jombart, T., Nedjati-Gilani, G., Nouvellet, P., Riley, S., et al., 2017. Key data for outbreak evaluation: building on the Ebola experience. Philosophical Transactions of the Royal Society B: Biological Sciences 372, 20160371.

Dong, E., Du, H., Gardner, L., 2020. An interactive web-based dashboard to track COVID-19 in real time. The Lancet Infectious Diseases 20, 533–534. doi:10.1016/S1473-3099(20)30120-1.

Elbe, S., Buckland-Merrett, G., 2017. Data, disease and diplomacy: GISAID’s innovative contribution to global health. Global Challenges 1, 33–46. doi:10.1002/gch2.1018.

Fernandez-Cassi, X., Scheidegger, A., Bänziger, C., Cariti, F., Corzon, A.T., Ganesanandamoorthy, P., Lemaitre, J.C., Ort, C., Julian, T.R., Kohn, T., 2021. Wastewater monitoring outperforms case numbers as a tool to track COVID-19 incidence dynamics when test positivity rates are high. Water Research 200, 117252.

Fox, S.J., Lachmann, M., Tec, M., Pasco, R., Woody, S., Du, Z., Wang, X., Ingle, T.A., Javan, E., Dahan, M., et al., 2022. Real-time pandemic surveillance using hospital admissions and mobility data. Proceedings of the National Academy of Sciences 119, e2111870119.

Greenwald, H.D., Kennedy, L.C., Hinkle, A., Whitney, O.N., Fan, V.B., Crits-Christoph, A., Harris-Lovett, S., Flamholz, A.I., Al-Shayeb, B., Liao, L.D., et al., 2021. Tools for interpretation of wastewater SARS-CoV-2 temporal and spatial trends demonstrated with data collected in the San Francisco Bay Area. Water Research 12, 100111.

Haver, A., Theijn, R., Grift, I.D., Raaijmakers, G., Poorter, E., Laros, J.F., van Dissel, J.T., Lodder, W.J., 2023. Regional reemergence of a SARS-CoV-2 Delta lineage amid an Omicron wave detected by wastewater sequencing. Scientific Reports 13, 17870.

Hodcroft, E., 2021. CoVariants: SARS-CoV-2 mutations and variants of interest. https://covariants.org.

Hoffmann, T., Alsing, J., 2023. Faecal shedding models for SARS-CoV-2 RNA among hospitalised patients and implications for wastewater-based epidemiology. Journal of the Royal Statistical Society Series C: Applied Statistics 72, 330–345.

Hopkins, L., Persse, D., Caton, K., Ensor, K., Schneider, R., McCall, C., Stadler, L.B., 2023. Citywide wastewater SARS-CoV-2 levels strongly correlated with multiple disease surveillance indicators and outcomes over three COVID-19 waves. Science of The Total Environment 855, 158967.

Huisman, J.S., Scire, J., Caduff, L., Fernandez-Cassi, X., Ganesanandamoorthy, P., Kull, A., Scheidegger, A., Stachler, E., Boehm, A.B., Hughes, B., et al., 2022. Wastewater-based estimation of the effective reproductive number of SARS-CoV-2. Environmental Health Perspectives 130, 057011.

Justman, J., Skalland, T., Moore, A., Amos, C.I., Marzinke, M.A., Zangeneh, S.Z., Kelley, C.F., Singer, R., Mayer, S., Hirsch-Moverman, Y., et al., 2024. Prevalence of SARS-CoV-2 infection among children and adults in 15 US communities, 2021. Emerging Infectious Diseases 30, 245.

Kaplan, E.H., Zulli, A., Sanchez, M., Peccia, J., 2022. Scaling SARS-CoV-2 wastewater concentrations to population estimates of infection. Scientific Reports 12, 3487.

Keeling, M.J., 2023. Patterns of reported infection and reinfection of SARS-CoV-2 in England. Journal of Theoretical Biology 556, 111299.

Larsen, D.A., Wigginton, K.R., 2020. Tracking COVID-19 with wastewater. Nature Biotechnology 38, 1151–1153.

Linton, N.M., Kobayashi, T., Yang, Y., Hayashi, K., Akhmetzhanov, A.R., Jung, S.m., Yuan, B., Kinoshita, R., Nishiura, H., 2020. Incubation period and other epidemiological characteristics of 2019 novel coronavirus infections with right truncation: a statistical analysis of publicly available case data. Journal of Clinical Medicine 9, 538.

Liu, J., Cai, Z., Gustafson, P., McDonald, D.J., 2024. rtestim: Time-varying reproduction number estimation with trend filtering. PLOS Computational Biology 20, e1012324.

Lobay, R., Srivastava, A., Tibshirani, R.J., McDonald, D.J., 2025. Incident COVID-19 infections before Omicron in the US. Epidemics 52, 100838.

Ma, K.C., 2023. Trends in laboratory-confirmed SARS-CoV-2 reinfections and associated hospitalizations and deaths among adults aged at least 18 years in 18 US jurisdictions, September 2021–December 2022. MMWR. Morbidity and Mortality Weekly Report 72.

McMahan, C.S., Self, S., Rennert, L., Kalbaugh, C., Kriebel, D., Graves, D., Colby, C., Deaver, J.A., Popat, S.C., Karanfil, T., et al., 2021. COVID-19 wastewater epidemiology: a model to estimate infected populations. The Lancet Planetary Health 5, e874–e881.

McManus, O., Christiansen, L.E., Nauta, M., Krogsgaard, L.W., Bahrenscheer, N.S., von Kappelgaard, L., Christiansen, T., Hansen, M., Hansen, N.C., Kahler, J., et al., 2023. Predicting COVID-19 incidence using wastewater surveillance data, Denmark, October 2021–June 2022. Emerging Infectious Diseases 29, 1589.

Medema, G., Heijnen, L., Elsinga, G., Italiaander, R., Brouwer, A., 2020. Presence of SARS-Coronavirus-2 RNA in sewage and correlation with reported COVID-19 prevalence in the early stage of the epidemic in the Netherlands. Environmental science & technology letters 7, 511–516.

Natarajan, A., Zlitni, S., Brooks, E.F., Vance, S.E., Dahlen, A., Hedlin, H., Park, R.M., Han, A., Schmidtke, D.T., Verma, R., et al., 2022. Gastrointestinal symptoms and fecal shedding of SARS-CoV-2 RNA suggest prolonged gastrointestinal infection. Med 3, 371–387.

Nourbakhsh, S., Fazil, A., Li, M., Mangat, C.S., Peterson, S.W., Daigle, J., Langner, S., Shurgold, J., D’Aoust, P., Delatolla, R., et al., 2022. A wastewater-based epidemic model for SARS-CoV-2 with application to three Canadian cities. Epidemics 39, 100560.

Okada, Y., Nishiura, H., 2024. Estimating the effective reproduction number of COVID-19 from population-wide wastewater data: An application in Kagawa, Japan. Infectious Disease Modelling 9, 645–656.

Pellis, L., Scarabel, F., Stage, H.B., Overton, C.E., Chappell, L.H., Fearon, E., Bennett, E., Lythgoe, K.A., House, T.A., Hall, I., et al., 2021. Challenges in control of COVID-19: Short doubling time and long delay to effect of interventions. Philosophical Transactions of the Royal Society B 376, 20200264.

Phan, T., Brozak, S., Pell, B., Ciupe, S.M., Ke, R., Ribeiro, R.M., Gitter, A., Mena, K.D., Perelson, A.S., Kuang, Y., et al., 2025. Post-recovery viral shedding shapes wastewater-based epidemiological inferences. Communications Medicine 5, 1–10.

Polo, D., Quintela-Baluja, M., Corbishley, A., Jones, D.L., Singer, A.C., Graham, D.W., Romalde, J.L., 2020. Making waves: wastewater-based epidemiology for COVID-19–approaches and challenges for surveillance and prediction. Water Research 186, 116404.

Prasek, S.M., Pepper, I.L., Innes, G.K., Slinski, S., Betancourt, W.Q., Foster, A.R., Yaglom, H.D., Porter, W.T., Engelthaler, D.M., Schmitz, B.W., 2023. Variant-specific SARS-CoV-2 shedding rates in wastewater. Science of The Total Environment 857, 159165.

Prasek, S.M., Pepper, I.L., Innes, G.K., Slinski, S., Ruedas, M., Sanchez, A., Brierley, P., Betancourt, W.Q., Stark, E.R., Foster, A.R., et al., 2022. Population level SARS-CoV-2 fecal shedding rates determined via wastewater-based epidemiology. Science of The Total Environment 838, 156535.

Rabe, A., Ravuri, S., Burnor, E., Steele, J.A., Kantor, R.S., Choi, S., Forman, S., Batjiaka, R., Jain, S., León, T.M., et al., 2023. Correlation between wastewater and COVID-19 case incidence rates in major California sewersheds across three variant periods. Journal of Water and Health 21, 1303–1317.

Ramdas, A., Tibshirani, R.J., 2016. Fast and flexible ADMM algorithms for trend filtering. Journal of Computational and Graphical Statistics 25, 839–858. doi:10.1080/10618600.2015.1054033.

Ravuri, S., Burnor, E., Routledge, I., Linton, N.M., Thakur, M., Boehm, A., Wolfe, M., Bischel, H.N., Naughton, C.C., Alexander, T.Y., et al., 2025. Estimating effective reproduction numbers using wastewater data from multiple sewersheds for SARS-CoV-2 in California counties. Epidemics 50, 100803.

Reinhart, A., Brooks, L., Jahja, M., Rumack, A., Tang, J., Agrawal, S., Al Saeed, W., Arnold, T., Basu, A., Bien, J., et al., 2021. An open repository of real-time COVID-19 indicators. Proceedings of the National Academy of Sciences 118, e2111452118.

Riley, S., Ainslie, K.E., Eales, O., Walters, C.E., Wang, H., Atchison, C., Fronterre, C., Diggle, P.J., Ashby, D., Donnelly, C.A., et al., 2021. Resurgence of SARS-CoV-2: detection by community viral surveillance. Science 372, 990–995.

Ruff, J., Zhang, Y., Kappel, M., Rathi, S., Watkins, K., Zhang, L., Lockett, C., 2022. Rapid increase in suspected SARS-CoV-2 reinfections, Clark County, Nevada, USA, December 2021. Emerging Infectious Diseases 28, 1977.

Russell, T.W., Townsley, H., Abbott, S., Hellewell, J., Carr, E.J., Chapman, L., Pung, R., Quilty, B.J., Hodgson, D., Fowler, A.S., et al., 2023. Within-host SARS-CoV-2 viral kinetics informed by complex life course exposures reveals different intrinsic properties of Omicron and Delta variants. medRxiv doi:10.1101/2023.05.17.23290105.

Schill, R., Nelson, K.L., Harris-Lovett, S., Kantor, R.S., 2023. The dynamic relationship between COVID-19 cases and SARS-CoV-2 wastewater concentrations across time and space: Considerations for model training data sets. Science of the Total Environment 871, 162069.

Soller, J., Jennings, W., Schoen, M., Boehm, A., Wigginton, K., Gonzalez, R., Graham, K.E., McBride, G., Kirby, A., Mattioli, M., 2022. Modeling infection from SARS-CoV-2 wastewater concentrations: Promise, limitations, and future directions. Journal of Water and Health 20, 1197–1211.

Tibshirani, R.J., 2014. Adaptive piecewise polynomial estimation via trend filtering. The Annals of Statistics 42, 285–323. doi:10.1214/13-AOS1189.

Tibshirani, R.J., 2022. Divided differences, falling factorials, and discrete splines: Another look at trend filtering and related problems. Foundations and Trends in Machine Learning 15, 694–846.

Washington State Department of Health, 2022. Reported COVID-19 reinfections in Washington State. https://doh.wa.gov/sites/default/files/2022-02/421-024-ReportedReinfections.pdf.

Watson, L.M., Plank, M.J., Armstrong, B.A., Chapman, J.R., Hewitt, J., Morris, H., Orsi, A., Bunce, M., Donnelly, C.A., Steyn, N., 2024. Jointly estimating epidemiological dynamics of Covid-19 from case and wastewater data in Aotearoa New Zealand. Communications Medicine 4, 143.

Xu, X., Wu, Y., Kummer, A.G., Zhao, Y., Hu, Z., Wang, Y., Liu, H., Ajelli, M., Yu, H., 2023. Assessing changes in incubation period, serial interval, and generation time of SARS-CoV-2 variants of concern: a systematic review and meta-analysis. BMC Medicine 21, 374.

Zhang, J., Litvinova, M., Wang, W., Wang, Y., Deng, X., Chen, X., Li, M., Zheng, W., Yi, L., Chen, X., et al., 2020. Evolving epidemiology and transmission dynamics of coronavirus disease 2019 outside Hubei province, China: a descriptive and modelling study. The Lancet Infectious Diseases 20, 793–802.

