## Supplement for "From Wastewater to Infection Estimates: Incident COVID-19 Infections during Omicron in the U.S."

#### List of Figures

A.1 Average effective genome copies per mL of wastewater by sampling week for the four US regions (Midwest, Northeast, South, and West) over January 2020 to March 2023. 3
A.2 Average effective genome copies per mL of wastewater by sampling week for North Carolina before (left) and after (right) applying the shrinkage procedure. . . . . . . 4
A.3 Estimated shedding rates for Delta and Omicron across 100 sets of Gaussian-process-based infections, with individual shedding rate estimates overlaid as points. . . . . . 8
A.4 Daily estimated $R_t$ using the median of infections for the seven states of California, Hawaii, Montana, North Carolina, Nevada, New York, and Washington. . . . . . . 9
A.5 Daily estimated $R_t$ using the 7-day average of cases for the seven states of California, Hawaii, Montana, North Carolina, Nevada, New York, and Washington. . . . . . . . 10
A.6 Daily estimated $R_t$ using the median of infections (blue) and 7-day average of cases (orange) for California, Hawaii, Montana, North Carolina, Nevada, New York, and Washington. These estimates are calculated using the tuning parameter that yields the optimal cross-validation score within one standard error, and are based on the lowest reported mean generation time and serial interval. . . . . . . . . . . . . . . . 11

A.7 Daily estimated $R_t$ using the median of infections (blue) and 7-day average of cases (orange) for California, Hawaii, Montana, North Carolina, Nevada, New York, and Washington. These estimates are calculated using the tuning parameter that yields the optimal cross-validation score within one standard error, and are based on the highest reported mean generation time and serial interval. . . . . . . . . . . . . . . 12
A.8 Daily relative change growth rate estimates for infections (blue) and the 7-day average of cases (orange), each calculated using a 7-day bandwidth. Reference lines at ±1% are provided in grey to indicate when growth rates surpass these bounds. . 13
A.9 Daily relative change growth rate estimates for infections (blue) and the 7-day average of cases (orange), each calculated using a 14-day bandwidth. Reference lines at ±1% are provided in grey to indicate when growth rates surpass these bounds. . 14
A.10 Daily relative change growth rate estimates for infections (blue) and the 7-day average of cases (orange), each calculated using a 21-day bandwidth. Reference lines at ±1% are provided in grey to indicate when growth rates surpass these bounds. . 15
A.11 Daily relative change growth rate estimates for infections (blue) and the 7-day average of cases (orange), each calculated using a 28-day bandwidth. Reference lines at ±1% are provided in grey to indicate when growth rates surpass these bounds. . 16
A.12 Estimated infections (assuming shedding profile from Watson et al. (2024)) and reported cases per 100,000 (7-day average, seven states) For visualization purposes, cases are transformed to match the scale of infections and the consecutive identical case values are omitted for each state, retaining only the first occurrence of each value. 17
A.13 Estimated infections (assuming shedding profile from Okada and Nishiura (2024)) and reported cases per 100,000 population averaged over seven days for seven states. For visualization purposes, cases are transformed to match the scale of infections and the consecutive identical case values are omitted for each state, retaining only the first occurrence of each value. . . . . . . . . . . . . . . . . . . . . . . . . . . 18
A.14 Estimated infections (assuming shedding profile from Huisman et al. (2022)) and reported cases per 100,000 population averaged over seven days for seven states. For visualization purposes, cases are transformed to match the scale of infections and the consecutive identical case values are omitted for each state, retaining only the first occurrence of each value. . . . . . . . . . . . . . . . . . . . . . . . . . . . . 19

### A Supplementary Methods and Results

#### A.1 Biobot analytics wastewater concentration data and constructing state-level estimates

Biobot Analytics provides the SARS-CoV-2 wastewater concentration data used in this study, normalized to the Pepper Mild Mottle Virus (PMMoV) fecal indicator to adjust for differences in dilution and population (Biobot Analytics Inc., 2020, 2022). Therefore, all subsequent references to wastewater concentration data refer to these PMMoV-normalized and laboratory protocol-adjusted SARS-CoV-2 concentrations.

Due to the lack of state-level wastewater concentration data, we opt for the county and regional estimates (for US census regions), that each provide up to one weekly population-weighted average of concentration in effective genome copies per mL of wastewater, with measurement dates set to Wednesday to reflect typical sample collection during the Monday to Friday work week.

To covert the county-level wastewater concentration measurements to state-level measurements for each reported day, we calculate a weighted average using the population of each contributing county as the weight. Additionally, any measurement that is negative is set to be missing. Now, we do not rely solely on these constructed state wastewater measurements because they are based on data from a relatively small subset of counties within each state.

To ensure stability for the shedding rate estimation, we use both county and regional data for the four US regions (Midwest, Northeast, South, and West), since the regional wastewater measurements are reliably reported every week of the period of interest (Figure A.1).

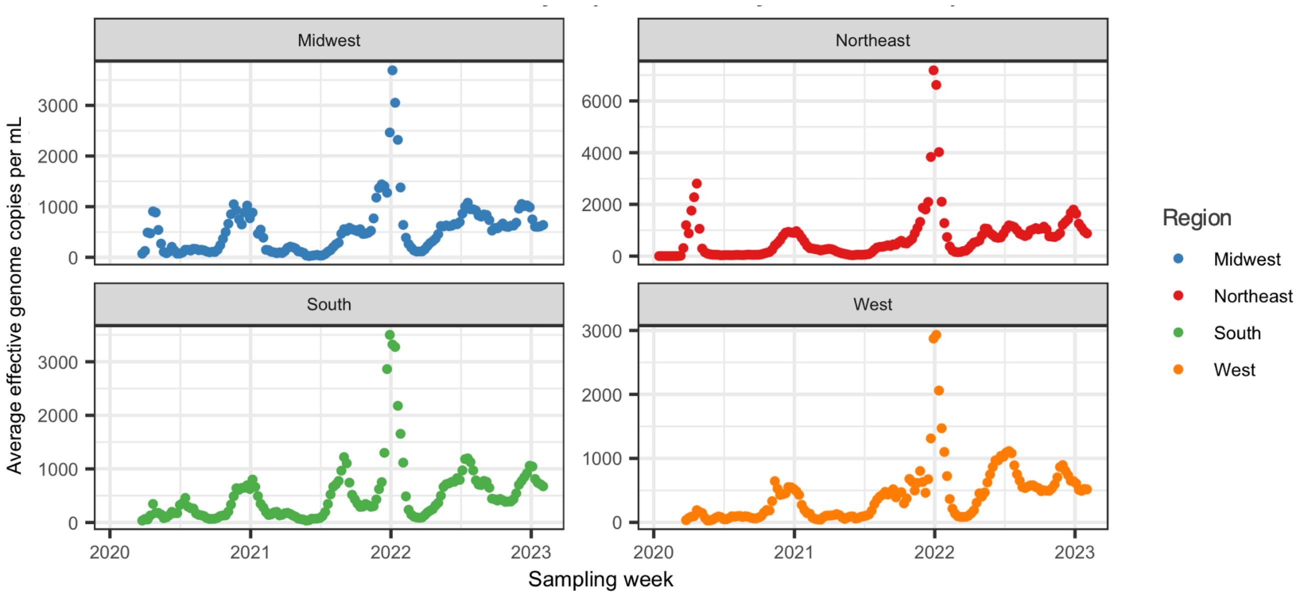

Figure A.1: Average effective genome copies per mL of wastewater by sampling week for the four US regions (Midwest, Northeast, South, and West) over January 2020 to March 2023.

During the period of Delta-Omicron overlap, when state wastewater data is missing, we fill these gaps by weighting the available state data by the fraction of the state population it represented and the corresponding regional data by the complement, and then take the weighted sum of these. If no state measurement was available, we take the regional value. In this way, we can construct a complete set of wastewater measurements for each state during this period. We call these the shifted wastewater concentration estimates.

Figure A.2 shows an example of the average effective genome copies per mL of wastewater for the state of North Carolina before and after applying this shrinkage procedure:

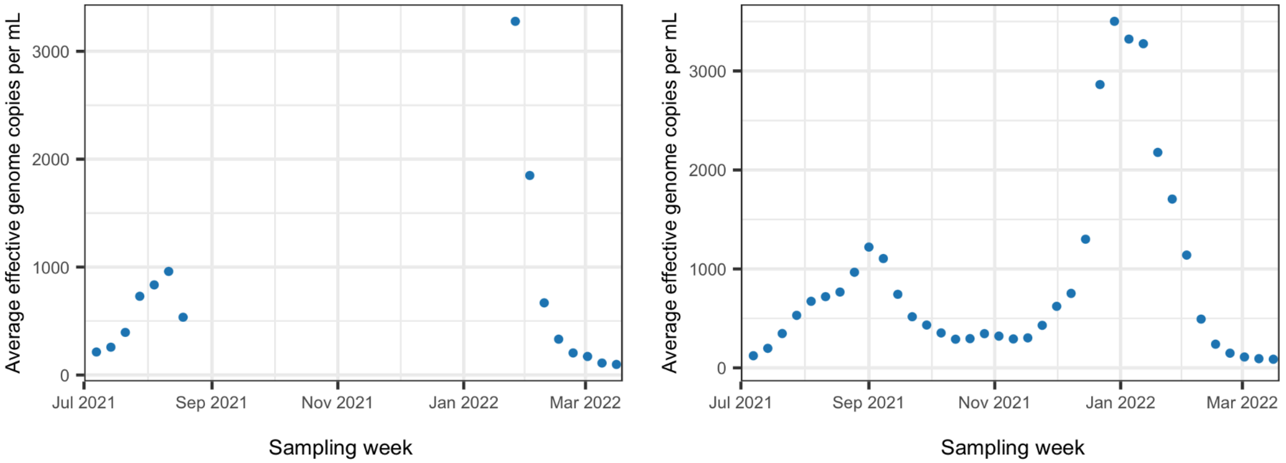

Figure A.2: Average effective genome copies per mL of wastewater by sampling week for North Carolina before (left) and after (right) applying the shrinkage procedure.

#### A.2 State-specific reinfection data and deconvolution for alignment

Accurate reinfection data, specific to each location, is important for inferring the total infection burden over Omicron, given the substantial increase in reinfections compared to previous variants and the clear limitations of relying solely on first infections (Ma, 2023; Ruff et al., 2022). We therefore compiled publicly available reinfection data from state health department websites. In total, such reinfection data was found for seven states: California, New York, Nevada, Washington, Hawaii, Minnesota, and North Carolina.[1] Of these states, only Minnesota and North Carolina provide daily reinfection data, while the others provide weekly data. Additionally, a concern is that the reinfection data are typically aligned to the date of positive specimen collection, but for accurate infection estimation, they should be aligned with the infection onset date. To push back the reported reinfections to their associated plausible date of infection onset (and to simultaneously deal with weekly reports), we apply a deconvolution technique that uses trend filtering, which is a nonparametric regression method known for its locally adaptive smoothness. Importantly, this adaptive smoothness allows for abrupt changes in the estimated reinfections, due to policy changes or other factors, while still promoting smoothness.

To specify the modelling setup, let $t = 1, \ldots, T_\ell$ denote the state-specific deconvolution period, depending on the start and end dates of that state's reinfection data. Define $y_{\ell,t}$ as the number of new reinfections reported by positive specimen date in state $\ell$ at time $t$ and the number of new reinfections with infection onset $k$ days earlier, $x_{t-k}$. To trace the timeline back and estimate reinfections by their infection onset, we use variant-specific delay distributions that reflect the time from infection to positive specimen collection. These are constructed by convolving variant-specific incubation period distributions with state-time-specific symptom-to-test delay distributions estimated from the CDC line list (Centers for Disease Control and Prevention, 2020), which gives delay distribution $\tau_{j,\ell,t-k}$ for variant $j$ in state $\ell$ and delay $k = 1, \ldots, K$. The most involved part is estimating the symptom-to-test delay distributions from the CDC line list. In brief, to estimate

[1]For the sources of the reinfection data, see California Department of Public Health (2021); Hawaii Department of Health (2022); Minnesota Department of Health (2020); New York State Department of Health (2021); North Carolina Department of Health and Human Services (2020); Ruff et al. (2022); Washington State Department of Health (2022).

these symptom-to-test delays, we extract onset and specimen collection dates from the individual cases in the line list and compute the empirical distribution of delays within a moving window centered around each date. We then fit a gamma distribution to the empirical delays via moment matching, and discretize the resulting density to form a probability mass function for the delay. This procedure follows the same general approach as described in Section 2.2 of Lobay et al. (2025). Estimating delay distributions by modelling observed delays with a probability distribution in this way is fairly standard in the COVID-19 and broader epidemiological literature (Abbott et al., 2020; Jahja et al., 2022; Miller et al., 2022; Park et al., 2024).

To deconvolve the reinfections by variant category (for the eight categories of Alpha, Beta, Gamma, Delta, Epsilon, Iota, Omicron, and Other), we weight the outcome (reinfection count) by the corresponding variant proportions in circulation that are also aligned to the date of positive specimen collection $v_{j,\ell,t}$. Then, the minimization problem for this daily reinfection data for variant $j$ in state $\ell$ is

$$\underset{\mathbf{x}}{\text{minimize}} \sum_t \left( v_{j,\ell,t} y_{\ell,t} - \sum_k \tau_{j,\ell,t-k} x_{t-k} \right)^2 + \lambda \| D^{(4)} \mathbf{x} \|_1,$$

where $\lambda$ is the regularization parameter that is selected using cross-validation and $D^{(4)}\mathbf{x} = x_t - 4x_{t-1} + 6x_{t-2} - 4x_{t-3} + x_{t-4}$, representing the fourth-order differences of $\mathbf{x}$.

To handle weekly reinfection data, we simply multiply the variant-specific convolution matrix by a matrix that encodes which days contribute to the weekly total. This matrix is then normalized so that for each time point $t$, for a fixed variant $j$ and location $\ell$, we have $\sum_k \tau_{j,\ell,t-k} = 1$.

As a result of deconvolving by variant, we estimate reinfections for each variant aligned to the date of infection onset. We use these updated reinfection estimates to refine the pre-Omicron inverse reporting ratios and corresponding infection estimates in the seven states. To do this, we apply a state-space model informed by two main antibody prevalence surveys in the U.S. (Centers for Disease Control and Prevention, 2021a,b), as detailed in previous work (refer to Sections 2.1–2.3 of Lobay et al., 2025). This antibody prevalence model treats weekly seroprevalence measurements as noisy observations of a latent seroprevalence process, which evolves over time based on new infections, reinfections, and antibody waning. Specifically, the model relates changes in latent seroprevalence to the product of deconvolved case counts and a time-varying inverse reporting ratio, accounting for the proportion of infections that are reinfections. The inverse reporting ratio itself is modelled as a smooth latent process. Thus, fitting this model yields updated estimates of both seroprevalence and inverse reporting ratios over time, which are then used to scale deconvolved cases to produce final infection estimates. Using this approach, we re-estimate infections from June 1, 2020 to February 21, 2022—the latter marking the earliest date at which reinfection reporting ends for a state: Minnesota lacks reinfection data beyond this point.

We apply the same methodology to estimate the infection onset dates for each state's new daily reported cases (Dong et al., 2020) over the entire period of interest (extending through February 1, 2023), allowing us to have deconvolved cases at our disposal to estimate infections during the Omicron era.

#### A.3 Gaussian process details

In the context of a Gaussian process, our goal is to simulate the time-varying inverse reporting ratios from a multivariate normal distribution specified for the inverse reporting ratio. To specify this multivariate normal distribution, we require the estimates of the inverse reporting ratio and the corresponding conditional covariance matrix for this state:

$$\mathbb{E}(a_1, \dots a_n \mid y_1, \dots, y_n)$$
$$\mathrm{Var}(a_1, \dots, a_n \mid y_1, \dots, y_n).$$

We can obtain the smoothed estimates of the inverse reporting ratio for each time, $\mathrm{E}(a_t \mid y_1, \dots, y_n)$ using Kalman filtering and smoothing. This is commonly implemented in R through the KFS function from the KFAS package (Helske, 2017). Unfortunately, we cannot obtain the correct covariance matrix from this. For the covariance matrices of the smoothed states, the KFS function gives a $n \times 4$ covariance matrix between the four states in $\alpha_t$ for each time under consideration: $\mathrm{Var}(\alpha_t \mid y_1, \dots, y_n)$. The problem is that this does not include the autocovariance between all of the elements of a particular state, $a_t$, which is what we need to have a complete conditional covariance matrix for $a_t$. To build the posterior variance using the results given by the KFAS functions, we use the matrix representation of the state space model as presented in Delle Monache and Petrella (2019), originally from Durbin and Koopman (2012). The following equations that specify the model structure are from Delle Monache and Petrella (2019) and form the basis for this approach:

$$y = B\alpha + \epsilon, \quad \epsilon \sim N(0, U),$$
$$\alpha = A(\alpha^* + R\eta), \quad \eta \sim N(0, V), \quad \alpha^* \sim N(a^*, P^*).$$

The observation equation components are

$$y = \begin{pmatrix} y_1 \\ \vdots \\ y_n \end{pmatrix}, \quad B = \begin{pmatrix} Z_1 & 0 & \cdots & 0 \\ 0 & Z_2 & \cdots & 0 \\ \vdots & \vdots & \ddots & \vdots \\ 0 & 0 & \cdots & Z_n \end{pmatrix}, \quad \epsilon = \begin{pmatrix} \epsilon_1 \\ \vdots \\ \epsilon_n \end{pmatrix}, \quad U = \begin{pmatrix} H_1 & 0 & \cdots & 0 \\ 0 & H_2 & \cdots & 0 \\ \vdots & \vdots & \ddots & \vdots \\ 0 & 0 & \cdots & H_n \end{pmatrix},$$

and the state equation components are

$$\alpha = \begin{pmatrix} \alpha_1 \\ \vdots \\ \alpha_n \end{pmatrix}, \quad A = \begin{pmatrix} I & 0 & \cdots & 0 \\ T_1 & I & \cdots & 0 \\ T_2T_1 & T_2 & \cdots & 0 \\ \vdots & \vdots & \ddots & \vdots \\ T_{n-1}\cdots T_1 & T_{n-1}T_{n-2} & \cdots & I \end{pmatrix}, \quad \alpha^* = \begin{pmatrix} \alpha_1 \\ 0 \\ \vdots \\ 0 \end{pmatrix}, \quad R = \begin{pmatrix} 0 & \cdots & 0 & \\ I & 0 & \cdots & 0 \\ 0 & I & \cdots & 0 \\ \vdots & \vdots & \ddots & \vdots \\ 0 & 0 & \cdots & I \end{pmatrix},$$

$$\eta = \begin{pmatrix} \eta_1 \\ \vdots \\ \eta_{n-1} \end{pmatrix}, \quad V = \begin{pmatrix} Q_1 & 0 & \cdots & 0 \\ 0 & Q_2 & \cdots & 0 \\ \vdots & \vdots & \ddots & \vdots \\ 0 & 0 & \cdots & Q_{n-1} \end{pmatrix}, \quad a^* = \begin{pmatrix} a_1 \\ 0 \\ \vdots \\ 0 \end{pmatrix}, \quad P^* = \begin{pmatrix} P_1 & 0 & \cdots & 0 \\ 0 & 0 & \cdots & 0 \\ \vdots & \vdots & \ddots & \vdots \\ 0 & 0 & \cdots & 0 \end{pmatrix},$$

and

$$G = \begin{pmatrix} P_1 & 0 & \cdots & 0 \\ 0 & Q_1 & \cdots & 0 \\ \vdots & \vdots & \ddots & \vdots \\ 0 & 0 & \cdots & Q_{n-1} \end{pmatrix}.$$

The dimensions of the main vectors and matrices are summarized in Table A.1.

| Variable | Dimensions |
|---|---|
| $y$ | $Nn \times 1$ |
| $\alpha, \alpha^*, a^*$ | $mn \times 1$ |
| $\eta$ | $m(n-1) \times 1$ |
| $B$ | $Nn \times mn$ |
| $U$ | $Nn \times Nn$ |
| $V$ | $m(n-1) \times m(n-1)$ |
| $R$ | $mn \times m(n-1)$ |
| $A$ | $mn \times mn$ |
| $P^*$ | $mn \times mn$ |
| $G$ | $mn \times mn$ |

Table A.1: Dimensions of the main vectors and matrices in the state space model.

For the observations and states, the joint distribution is

$$\begin{pmatrix} \alpha \\ y \end{pmatrix} \sim N\left(\begin{pmatrix} \mu_\alpha \\ \mu_y \end{pmatrix}, \begin{pmatrix} \Sigma_{\alpha\alpha} & \Sigma_{\alpha y} \\ \Sigma_{\alpha y}^\top & \Sigma_{yy} \end{pmatrix}\right),$$

where

$$\begin{aligned} \mu_\alpha &= Aa^*, \\ \Sigma_{\alpha\alpha} &= AGA^\top, \\ \Sigma_{\alpha y} &= \Sigma_{\alpha\alpha} B', \\ \mu_y &= B\mu_\alpha, \\ \Sigma_{yy} &= B\Sigma_{\alpha\alpha}B^\top + U. \end{aligned}$$

By an elementary result in multivariate regression theory, the conditional distribution of $\alpha$ given $y$ is

$$\begin{aligned} \alpha|y &\sim N(\mu_{\alpha|y}, \Sigma_{\alpha\alpha|y}), \\ \mu_{\alpha|y} &= \mu_\alpha + \Sigma_{\alpha\alpha}B^\top(B\Sigma_{\alpha\alpha}B^\top + U)^{-1}(y - \mu_y), \\ \Sigma_{\alpha\alpha|y} &= \Sigma_{\alpha\alpha} - \Sigma_{\alpha\alpha}B^\top(B\Sigma_{\alpha\alpha}B^\top + U)^{-1}B\Sigma_{\alpha\alpha}, \end{aligned}$$

where

$$\mu_{\alpha|y} = \begin{pmatrix} a_{1|n} \\ \vdots \\ a_{n|n} \end{pmatrix}, \quad \Sigma_{\alpha\alpha|y} = \begin{pmatrix} P_{1|n} & \cdots & P_{1n|n} \\ \vdots & \ddots & \vdots \\ P_{n1|n} & \cdots & P_{n|n} \end{pmatrix}.$$

The conditional covariance matrix $\Sigma_{\alpha\alpha|y}$ is our target. Observe that the computation of this matrix involves operations amongst matrices and so can become computationally inefficient when large matrices are involved. Even so, we opt for its direct computation because we consider the estimation of the inverse reporting ratios using the state space model and repeated estimation under the Gaussian process to be two separate, though related procedures. The former should occur before the latter and is essential to the general approach we use to estimate infections (this was a fundamental part of the procedure to obtain the inverse reporting ratios and, hence, infections prior to Omicron). Thus, after performing the former, it is natural to use its matrices for further computations, including, but not limited to computing the conditional covariance matrix.

For estimating inverse reporting ratios over the Omicron period, simulating once from the multivariate normal distribution will give us one daily inverse reporting ratio per week over the range of considered dates, which is determined by the seroprevalence measurements. After, we proceed to fill-in one daily measurement per week outside the range in both directions using forward backward extrapolation. Then, we linearly interpolate to obtain one inverse reporting ratio for every day of the week. Ultimately, we obtain one complete set of new inverse reporting ratios between the pre-specified start date to end date that is fixed for all US states under study.

#### A.4 Estimated shedding rates for Delta and Omicron

Figure A.3 shows the estimated shedding rates for Delta and Omicron obtained by fitting the linear shedding model across 100 sets of Gaussian-process-based infection estimates. Note that each point represents the shedding rate estimates from a single fit and is accompanied by a box plot to indicate their center and spread.

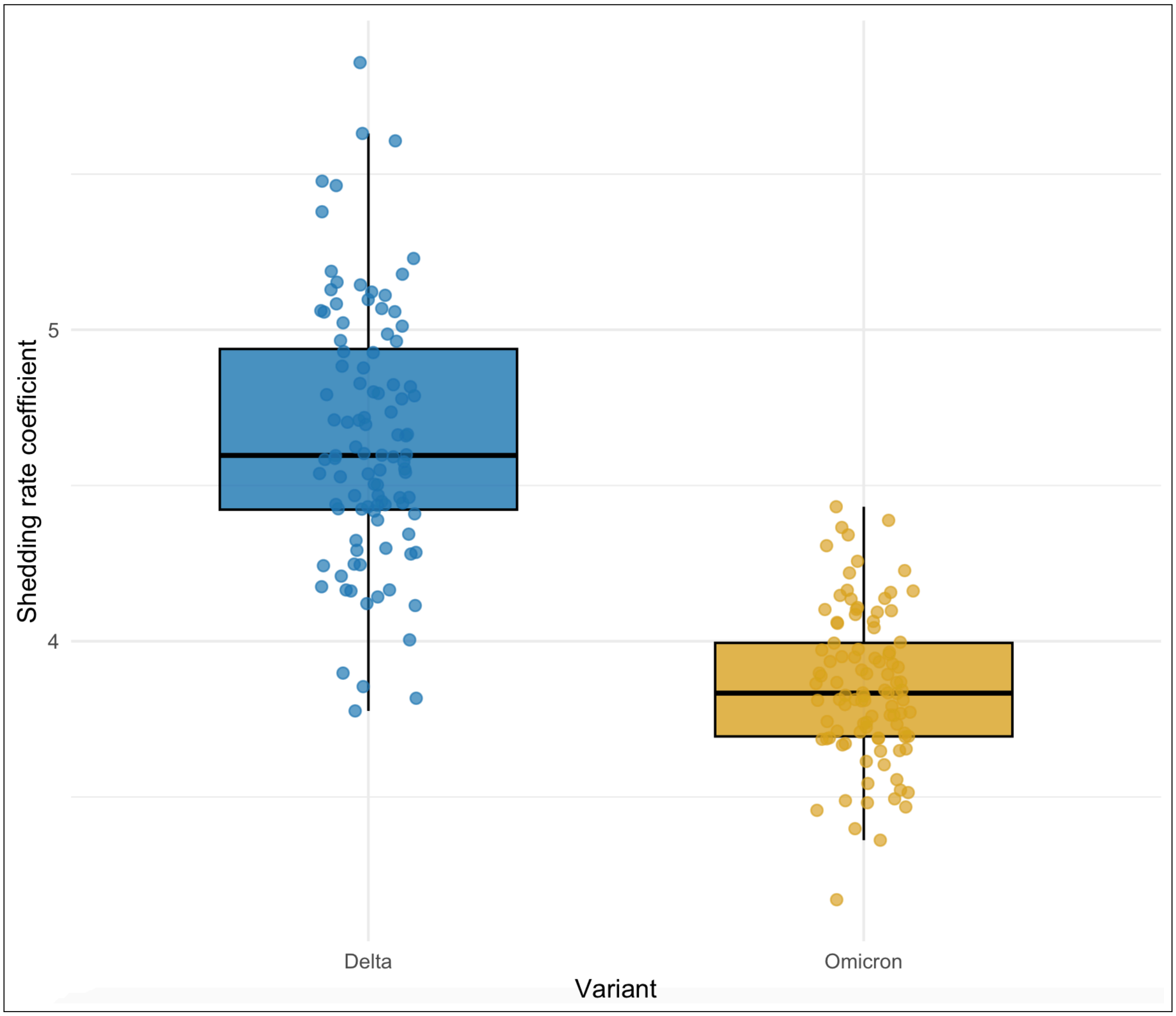

Figure A.3: Estimated shedding rates for Delta and Omicron across 100 sets of Gaussian-process-based infections, with individual shedding rate estimates overlaid as points.

#### A.5 $R_t$ cross-validation results

Figures A.4 and A.5 show the daily estimated $R_t$, which was derived using the median of infections and the 7-day average of cases across twenty different tuning parameters selected during cross-validation to identify the optimal parameter for each state.

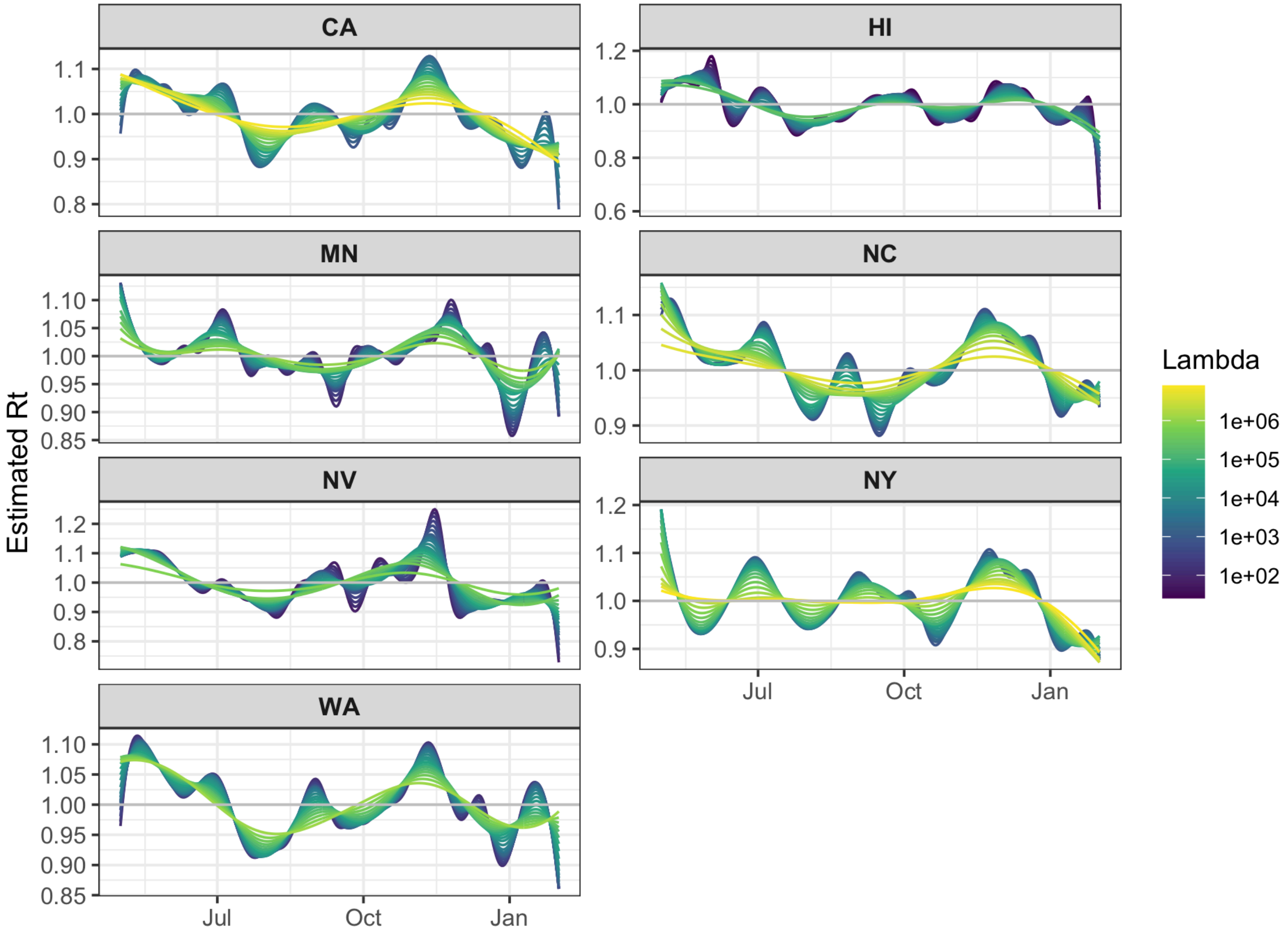

Figure A.4: Daily estimated $R_t$ using the median of infections for the seven states of California, Hawaii, Minnesota, North Carolina, Nevada, New York, and Washington.

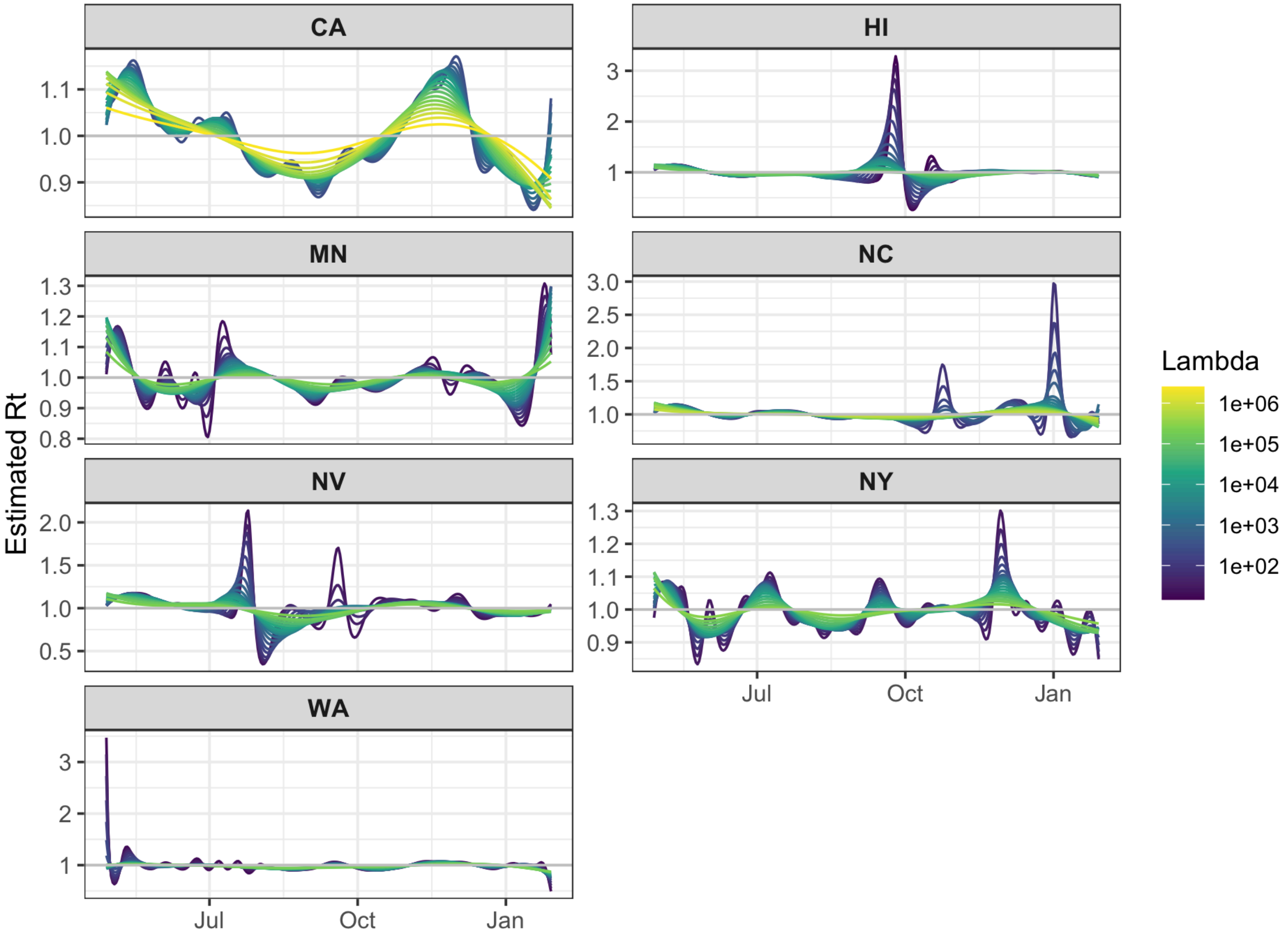

Figure A.5: Daily estimated $R_t$ using the 7-day average of cases for the seven states of California, Hawaii, Minnesota, North Carolina, Nevada, New York, and Washington.

#### A.6 Sensitivity of $R_t$ to varying generation times and serial intervals

In this section, we report the results of the sensitivity analysis using the lowest and highest mean generation times and serial intervals for Omicron, as reported in the meta-analysis by Xu et al. (2023). We briefly examine the plots to assess the similarity of the resulting $R_t$ estimates.

Figures A.6 and A.7 show the daily $R_t$ estimates, using the lowest and highest reported generation times and serial intervals, respectively. The most striking difference between these plots is that the case-based $R_t$ estimates become increasingly volatile as the mean and standard deviation of the serial interval increase (from a mean of 2.02 days with SD of 2.07 to 4.8 days with SD of 3), while the infection-based $R_t$ estimates remain relatively stable, increasing only slightly as the generation time increases (from a mean of 2.36 days with a SD of 3.7 to 3.59 days with the same SD). The infection-based $R_t$ results tend to be similar in pattern and peak placement, suggesting that they are fairly robust to variations in these parameters, while the case-based estimates clearly lack any such robustness.

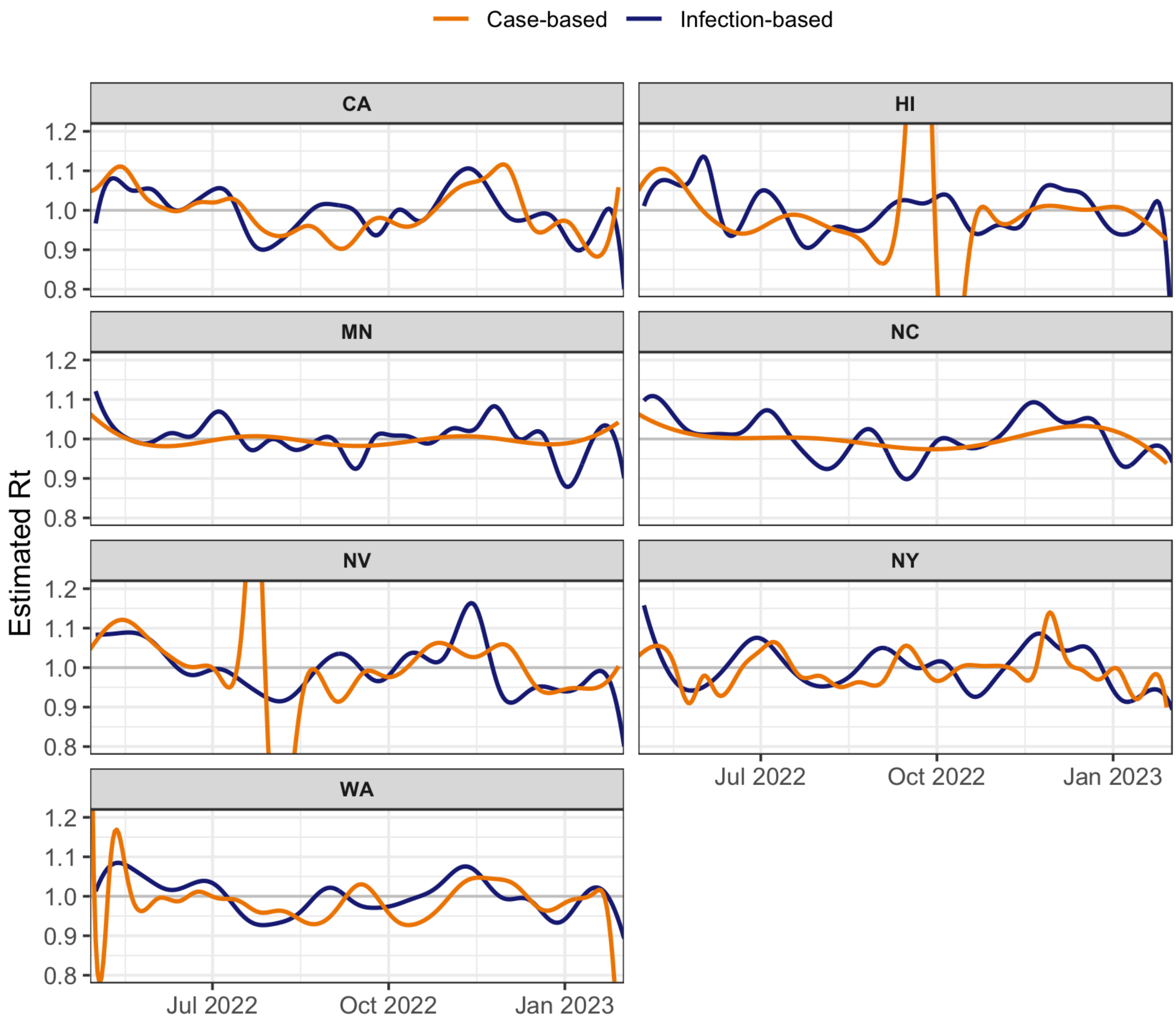

Figure A.6: Daily estimated $R_t$ using the median of infections (blue) and 7-day average of cases (orange) for California, Hawaii, Minnesota, North Carolina, Nevada, New York, and Washington. These estimates are calculated using the tuning parameter that yields the optimal cross-validation score within one standard error, and are based on the lowest reported mean generation time and serial interval.

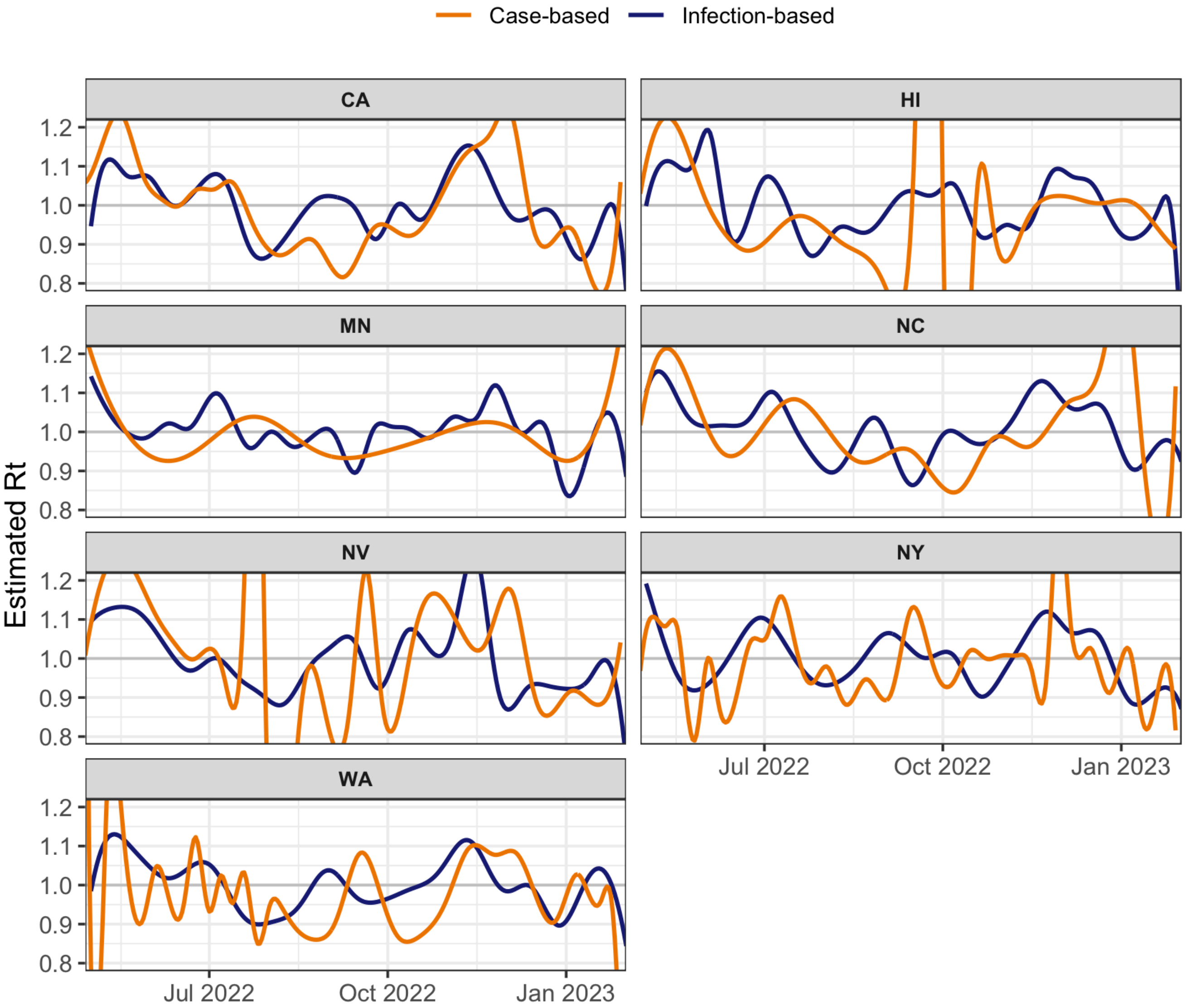

Figure A.7: Daily estimated $R_t$ using the median of infections (blue) and 7-day average of cases (orange) for California, Hawaii, Minnesota, North Carolina, Nevada, New York, and Washington. These estimates are calculated using the tuning parameter that yields the optimal cross-validation score within one standard error, and are based on the highest reported mean generation time and serial interval.

#### A.7 Sensitivity of growth rate estimates to varying window sizes

To evaluate the impact of bandwidth choice on the growth rate estimation, we present a sensitivity analysis that considers alternative bandwidths of 7, 14, 21, and 28 days (including the reference 14-day window). Figures A.8 to A.11 show similar overall trends across the bandwidths, though it is apparent that as the bandwidth increases, the growth rates become smoother, particularly for the case-based estimates that are more prone to noise and reporting artifacts.

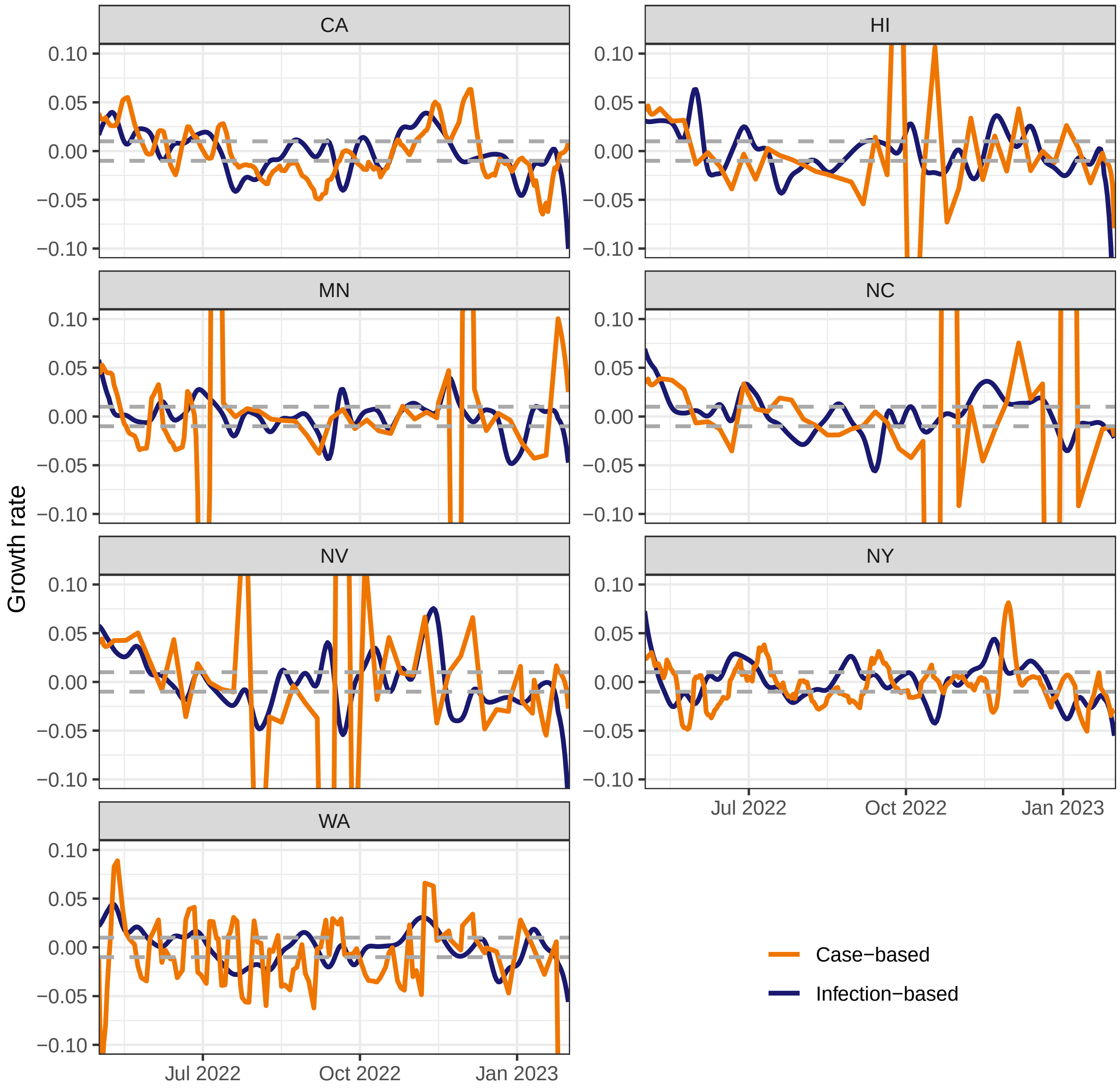

Figure A.8: Daily relative change growth rate estimates for infections (blue) and the 7-day average of cases (orange), each calculated using a 7-day bandwidth. Reference lines at ±1% are provided in grey to indicate when growth rates surpass these bounds.

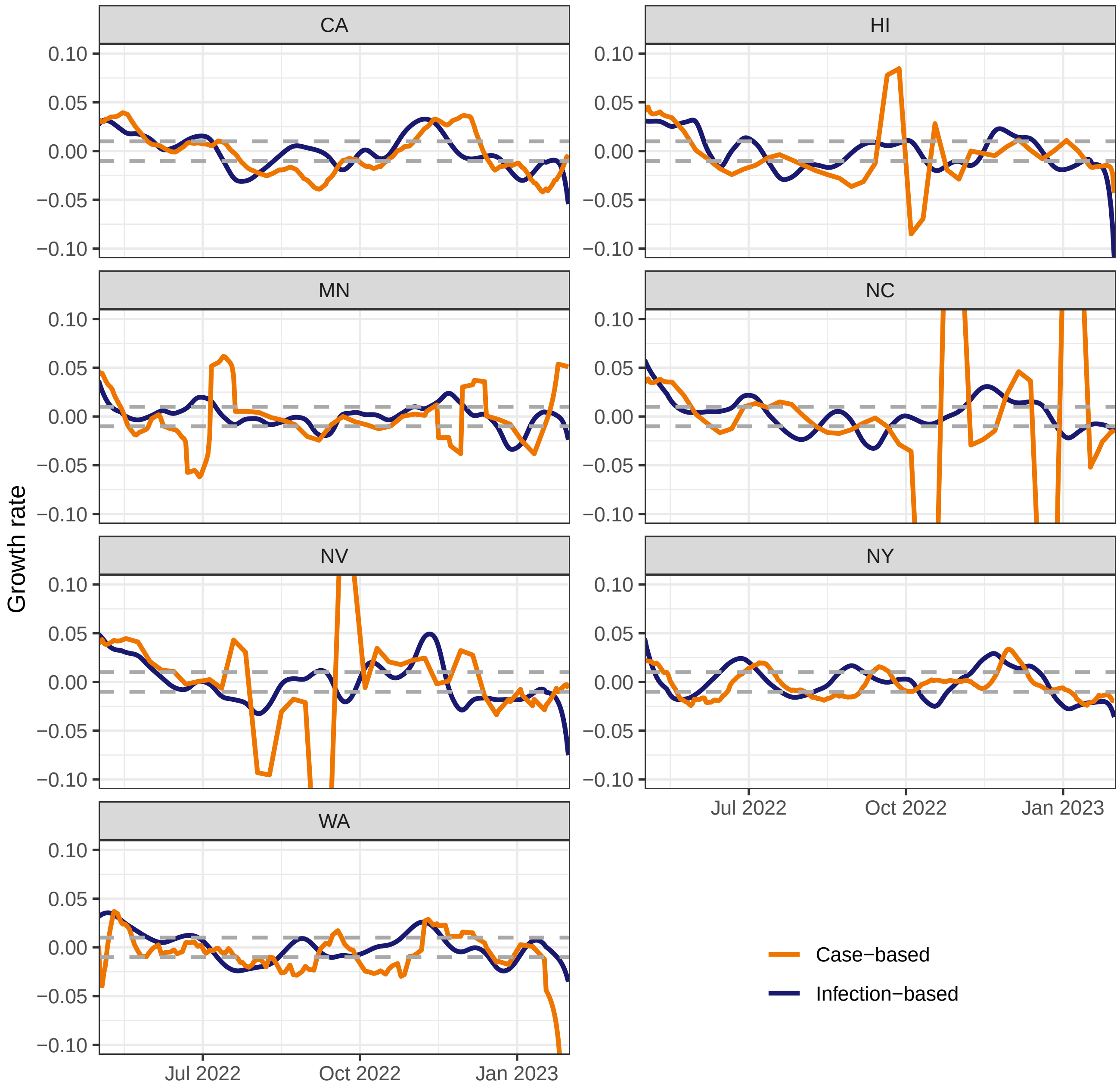

Figure A.9: Daily relative change growth rate estimates for infections (blue) and the 7-day average of cases (orange), each calculated using a 14-day bandwidth. Reference lines at ±1% are provided in grey to indicate when growth rates surpass these bounds.

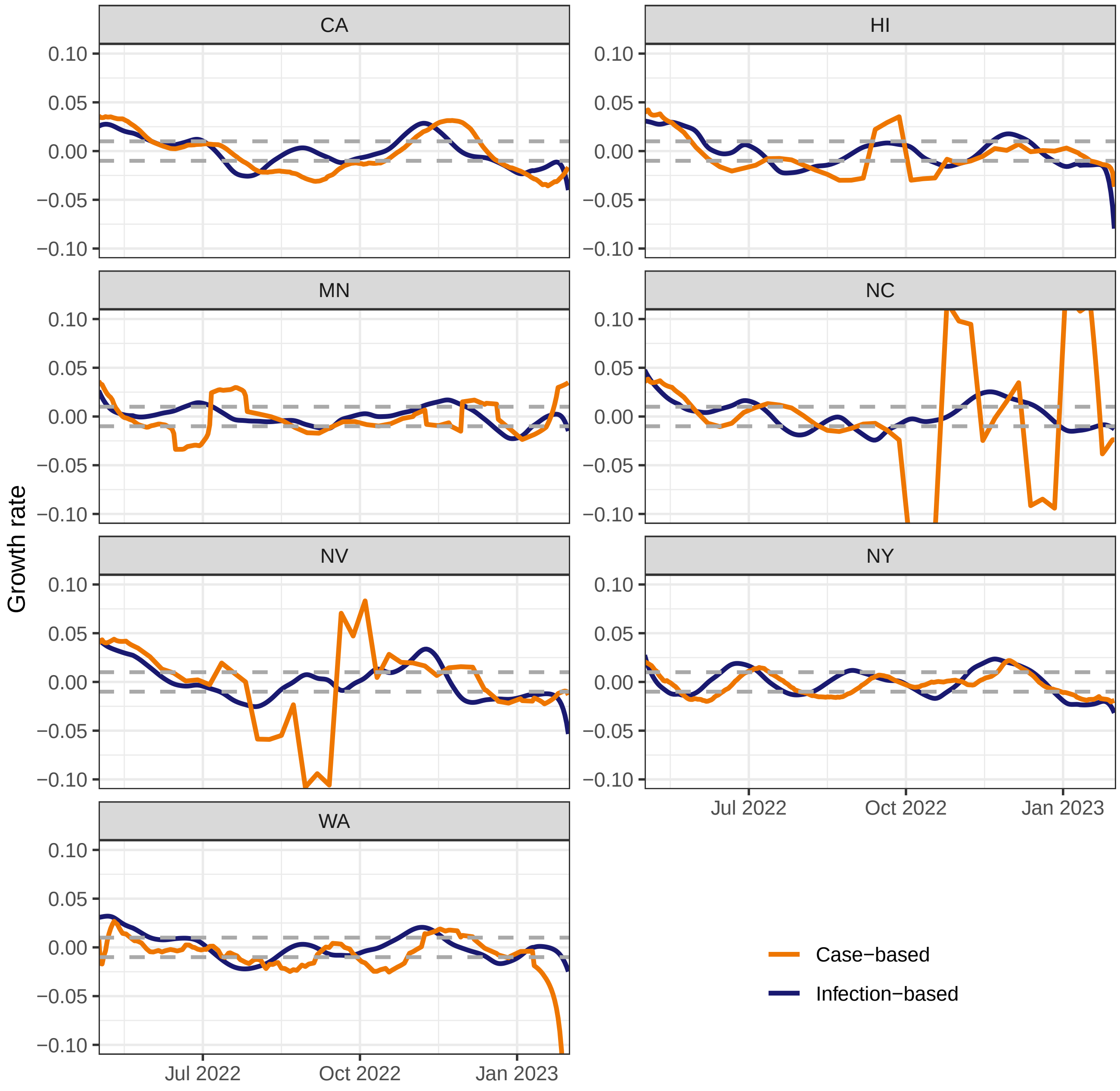

Figure A.10: Daily relative change growth rate estimates for infections (blue) and the 7-day average of cases (orange), each calculated using a 21-day bandwidth. Reference lines at ±1% are provided in grey to indicate when growth rates surpass these bounds.

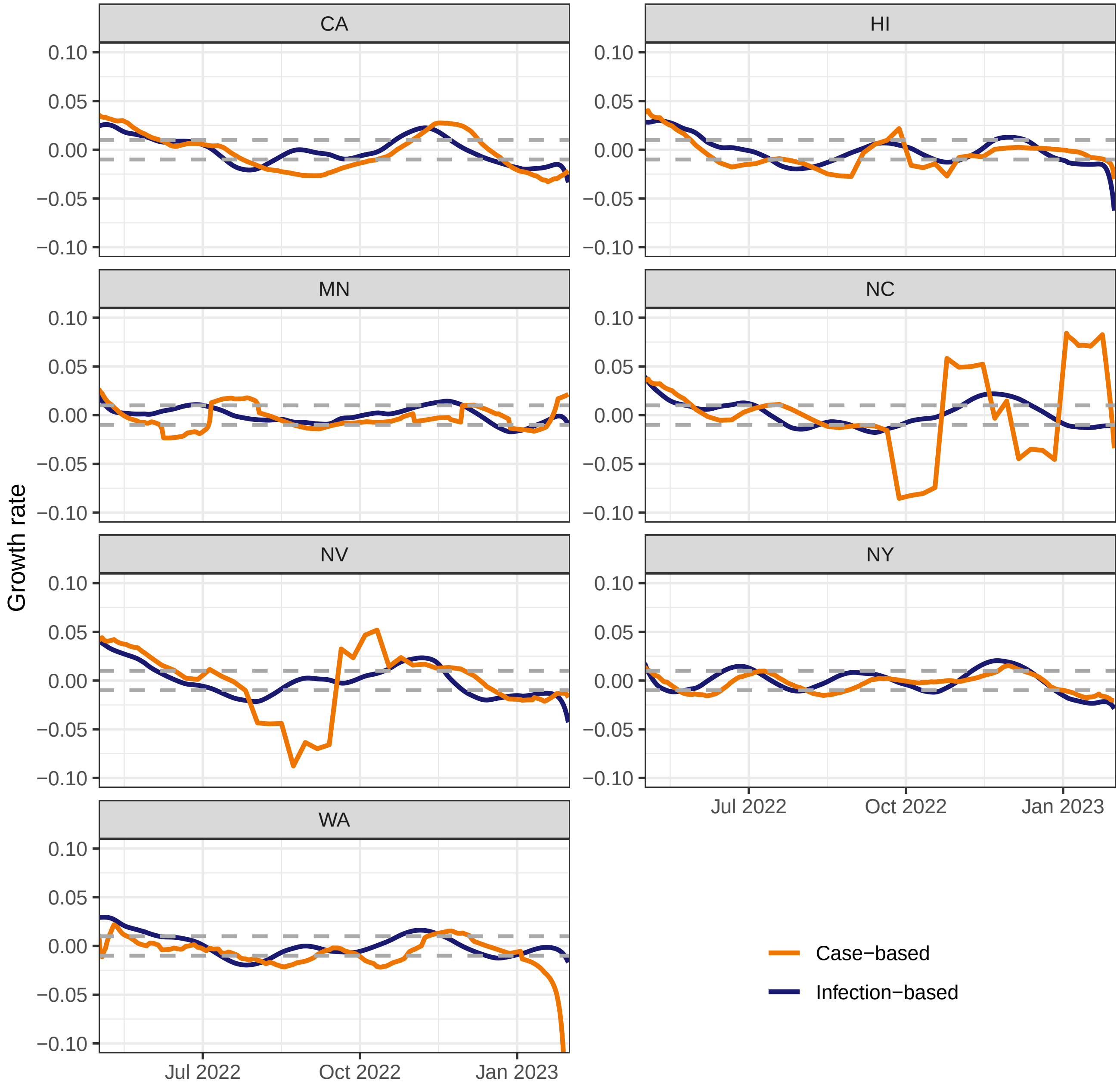

Figure A.11: Daily relative change growth rate estimates for infections (blue) and the 7-day average of cases (orange), each calculated using a 28-day bandwidth. Reference lines at ±1% are provided in grey to indicate when growth rates surpass these bounds.

#### A.8 Sensitivity of infection estimates to assumed shedding profile

Figures A.12 to A.14 show the estimated infection trajectories per 100,000 assuming that the shedding profiles are as in Watson et al. (2024) (original), Okada and Nishiura (2024), and Huisman et al. (2022), respectively. Overall, the magnitude, shape, and spread of the resulting infection trajectories are comparable and quite difficult to distinguish visually for Figure A.12 and Figure A.13 (save for the end estimates, which differ due to apparent boundary effects). This is owing to their similar Omicron shedding rates, with Watson et al. (2024) having a mean of 3.85 (SD 0.24) and

Okada and Nishiura (2024) having a mean of 4.01 (SD 0.25), which roughly scale the inverse reporting ratio estimates and, hence, inversely affect infection estimates during the Omicron era. Huisman et al. (2022) has the lowest mean shedding rate of 3.72 (SD 0.26), which results in slightly higher infection estimates than the others, though the same overall patterns persist. This relative comparability suggests that the infection estimates are reasonably robust to variations in the assumed shedding profile distribution.

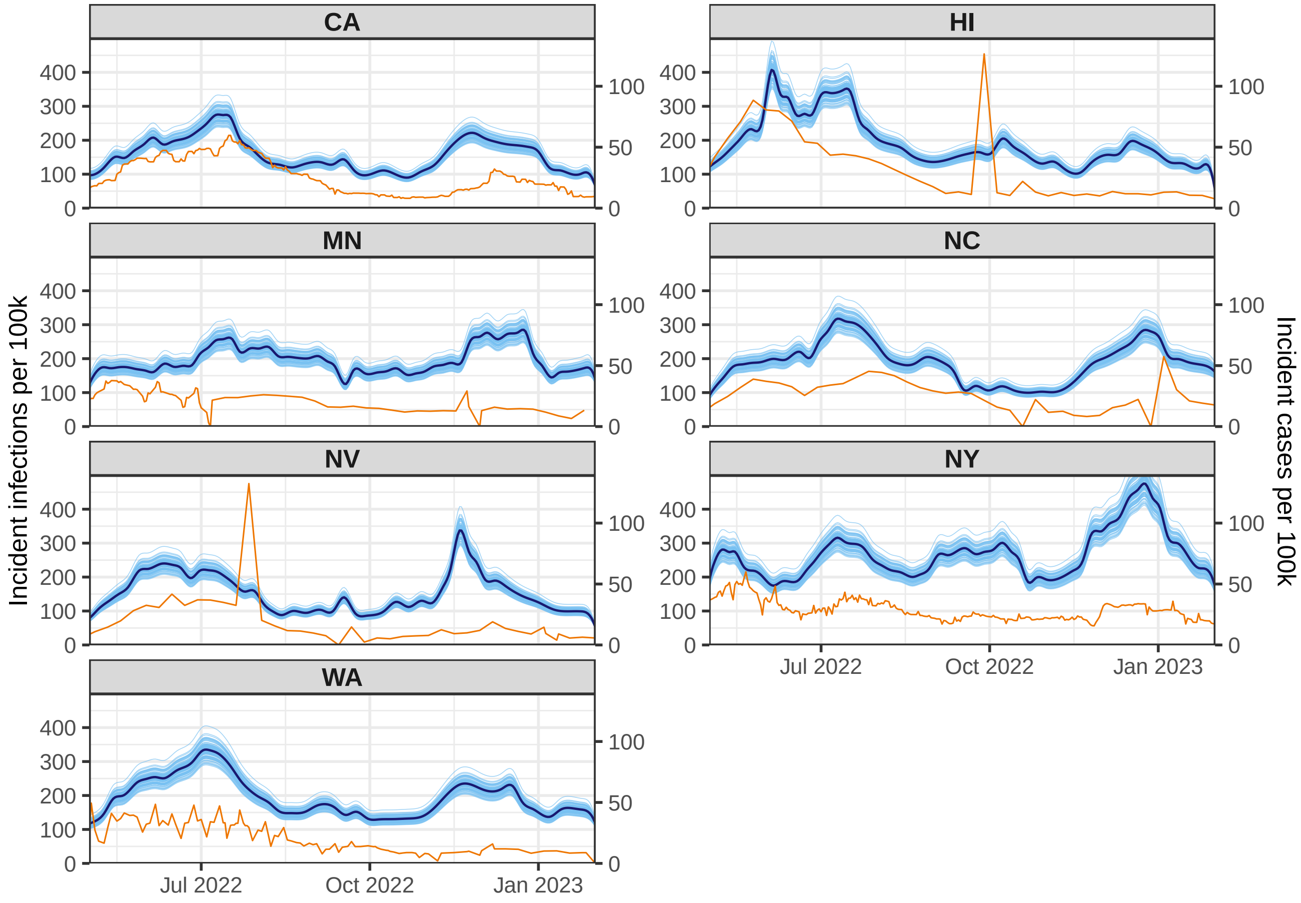

Figure A.12: Estimated infections (assuming shedding profile from Watson et al. (2024)) and reported cases per 100,000 (7-day average, seven states) For visualization purposes, cases are transformed to match the scale of infections and the consecutive identical case values are omitted for each state, retaining only the first occurrence of each value.

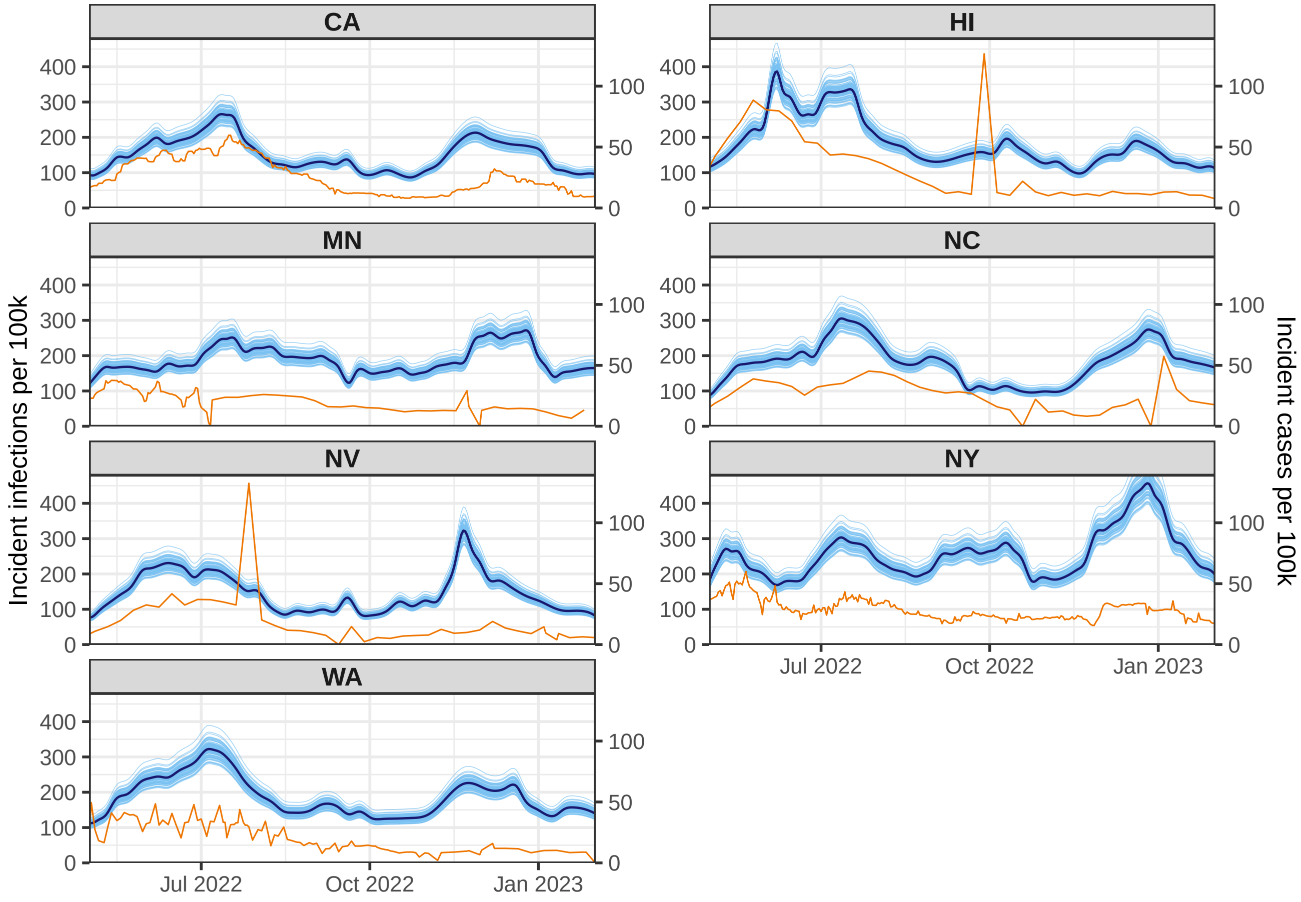

Figure A.13: Estimated infections (assuming shedding profile from Okada and Nishiura (2024)) and reported cases per 100,000 population averaged over seven days for seven states. For visualization purposes, cases are transformed to match the scale of infections and the consecutive identical case values are omitted for each state, retaining only the first occurrence of each value.

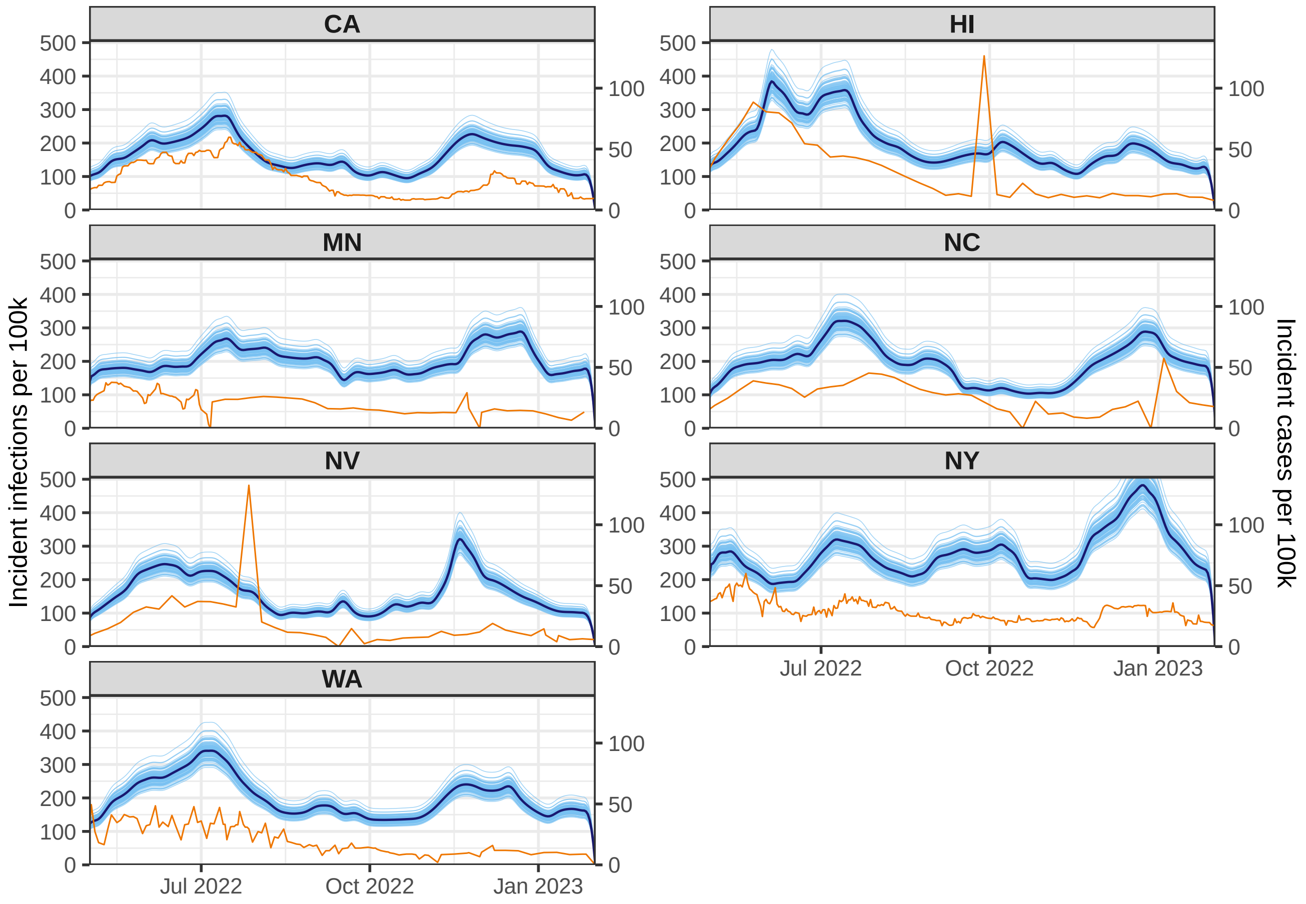

Figure A.14: Estimated infections (assuming shedding profile from Huisman et al. (2022)) and reported cases per 100,000 population averaged over seven days for seven states. For visualization purposes, cases are transformed to match the scale of infections and the consecutive identical case values are omitted for each state, retaining only the first occurrence of each value.

### References

Abbott, S., Hellewell, J., Thompson, R. N., Sherratt, K., Gibbs, H. P., Bosse, N. I., Munday, J. D., Meakin, S., Doughty, E. L., Chun, J. Y., et al., 2020. Estimating the time-varying reproduction number of SARS-CoV-2 using national and subnational case counts. Wellcome Open Research 5 (112), 112.

Biobot Analytics Inc., 2020. Biobot web dashboard – COVID-19 wastewater monitoring in the US. https://biobot.io/data/covid-19.

Biobot Analytics Inc., 2022. Effective concentration: a future-proof approach to characterizing pathogen concentrations in wastewater. https://web.archive.org/web/20230406191212/https://biobot.io/wp-content/uploads/2022/02/2022-01-White-paper-Effective-concentration.pdf.

California Department of Public Health, 2021. Reinfections account for 1 in 7 new COVID cases in July. https://www.mercurynews.com/2022/07/31/covid-1-in-7-new-california-cases-this-month-are-reinfections/.

Centers for Disease Control and Prevention, 2020. COVID-19 case surveillance restricted access detailed data. https://data.cdc.gov/Case-Surveillance/COVID-19-Case-Surveillance-Restricted-Access-Detai/mbd7-r32t.

Centers for Disease Control and Prevention, 2021a. 2020-2021 nationwide blood donor seroprevalence survey infection-induced seroprevalence estimates. https://data.cdc.gov/Laboratory-Surveillance/2020-2021-Nationwide-Blood-Donor-Seroprevalence-Su/mtc3-kq6r.

Centers for Disease Control and Prevention, 2021b. Nationwide commercial laboratory seroprevalence survey. https://data.cdc.gov/Laboratory-Surveillance/Nationwide-Commercial-Laboratory-Seroprevalence-Su/d2tw-32xv.

Delle Monache, D., Petrella, I., 2019. Efficient matrix approach for classical inference in state space models. Economics Letters 181, 22–27.

Dong, E., Du, H., Gardner, L., 2020. An interactive web-based dashboard to track COVID-19 in real time. The Lancet Infectious Diseases 20 (5), 533–534.

Durbin, J., Koopman, S. J., 2012. Time Series Analysis by State Space Methods. Vol. 38. OUP Oxford.

Hawaii Department of Health, 2022. COVID-19 reinfection data. https://health.hawaii.gov/coronavirusdisease2019/files/2022/09/reinfection_report_2022-09-28.pdf.

Helske, J., 2017. KFAS: Exponential family state space models in R. Journal of Statistical Software 78, 1–39.

Huisman, J. S., Scire, J., Caduff, L., Fernandez-Cassi, X., Ganesanandamoorthy, P., Kull, A., Scheidegger, A., Stachler, E., Boehm, A. B., Hughes, B., et al., 2022. Wastewater-based estimation of the effective reproductive number of SARS-CoV-2. Environmental Health Perspectives 130 (5), 057011.

Jahja, M., Chin, A., Tibshirani, R. J., 2022. Real-time estimation of COVID-19 infections: Deconvolution and sensor fusion. Statistical Science 37 (2), 207–228.

Lobay, R., Srivastava, A., Tibshirani, R. J., McDonald, D. J., 2025. Incident COVID-19 infections before Omicron in the US. Epidemics 52, 100838.

Ma, K. C., 2023. Trends in laboratory-confirmed SARS-CoV-2 reinfections and associated hospitalizations and deaths among adults aged at least 18 years in 18 US jurisdictions, September 2021–December 2022. MMWR. Morbidity and Mortality Weekly Report 72.

Miller, A. C., Hannah, L. A., Futoma, J., Foti, N. J., Fox, E. B., D'Amour, A., Sandler, M., Saurous, R. A., Lewnard, J. A., 2022. Statistical deconvolution for inference of infection time series. Epidemiology 33 (4), 470–479.

Minnesota Department of Health, 2020. Reinfections by date specimen collected data table. https://web.archive.org/web/20220225203802/https://www.health.state.mn.us/diseases/coronavirus/situation.html#casesro1.

New York State Department of Health, 2021. COVID-19 reinfection data. https://coronavirus.health.ny.gov/covid-19-reinfection-data.

North Carolina Department of Health and Human Services, 2020. North Carolina (archive) COVID-19 cases and deaths dashboard. https://covid19.ncdhhs.gov/dashboard/cases-and-deaths.

Okada, Y., Nishiura, H., 2024. Estimating the effective reproduction number of COVID-19 from population-wide wastewater data: An application in Kagawa, Japan. Infectious Disease Modelling 9 (3), 645–656.

Park, S. W., Akhmetzhanov, A. R., Charniga, K., Cori, A., Davies, N. G., Dushoff, J., Funk, S., et al., 2024. Estimating epidemiological delay distributions for infectious diseases. medRxiv.

Ruff, J., Zhang, Y., Kappel, M., Rathi, S., Watkins, K., Zhang, L., Lockett, C., 2022. Rapid increase in suspected SARS-CoV-2 reinfections, Clark County, Nevada, USA, December 2021. Emerging Infectious Diseases 28 (10), 1977.

Washington State Department of Health, 2022. Reported COVID-19 reinfections in Washington State. https://doh.wa.gov/sites/default/files/2022-02/421-024-ReportedReinfections.pdf.

Watson, L. M., Plank, M. J., Armstrong, B. A., Chapman, J. R., Hewitt, J., Morris, H., Orsi, A., Bunce, M., Donnelly, C. A., Steyn, N., 2024. Jointly estimating epidemiological dynamics of Covid-19 from case and wastewater data in Aotearoa New Zealand. Communications Medicine 4 (1), 143.

Xu, X., Wu, Y., Kummer, A. G., Zhao, Y., Hu, Z., Wang, Y., Liu, H., Ajelli, M., Yu, H., 2023. Assessing changes in incubation period, serial interval, and generation time of SARS-CoV-2 variants of concern: a systematic review and meta-analysis. BMC Medicine 21 (1), 374.
